# Integrative Modeling of the Spread of Serious Infectious Diseases and Corresponding Wastewater Dynamics

**DOI:** 10.1101/2024.11.10.24317057

**Authors:** Nina Schmid, Julia Bicker, Andreas F. Hofmann, Karina Wallrafen-Sam, David Kerkmann, Andreas Wieser, Martin J. Kühn, Jan Hasenauer

## Abstract

The COVID-19 pandemic has emphasized the critical need for accurate disease modeling to inform public health interventions. Traditional reliance on confirmed infection data is often hindered by reporting delays and under-reporting, while widespread antigen and antibody testing can be costly and impractical. Wastewater-based surveillance offers a promising alternative by detecting viral concentrations from fecal shedding, potentially providing a more accurate estimate of true infection prevalence. However, challenges remain in optimizing sampling protocols, locations, and normalization strategies, particularly in accounting for environmental factors like precipitation.

We present an integrative model that simulates the spread of serious infectious diseases by linking detailed infection dynamics with wastewater processes through viral shedding curves. Through comprehensive simulations, we examine how virus characteristics, precipitation events, measurement protocols, and normalization strategies affect the relationship between infection dynamics and wastewater measurements. Our findings reveal a complex relationship between disease prevalence and corresponding wastewater concentrations, with key variability sources including upstream sampling locations, continuous rainfall, and rapid viral decay. Notably, we find that flow rate normalization can be unreliable when rainwater infiltrates sewer systems. Despite these challenges, our study demonstrates that wastewater-based surveillance data can serve as a leading indicator of disease prevalence, predicting outbreak peaks before they occur. The proposed integrative model can thus be used to optimize wastewater-based surveillance, enhancing its utility for public health monitoring.

**Graphical Abstract:** 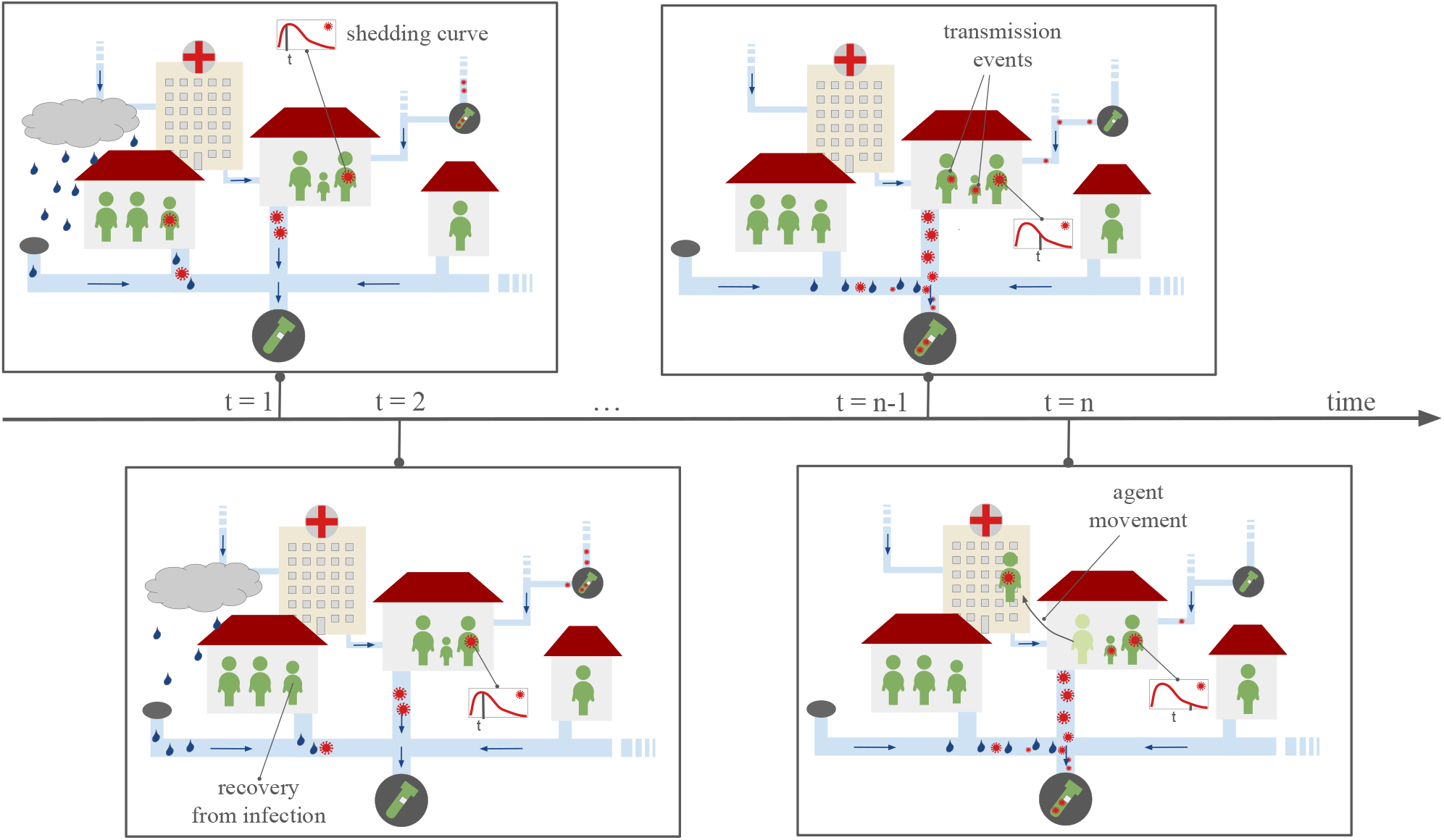

**Highlights:**

- Integration of infection and wastewater models to simulate disease spread.
- Identification of factors affecting wastewater measurements.
- Illustration of ability of wastewater-based surveillance to predict outbreaks before case reporting.
- Demonstration of unreliability of flow rate normalization in case of rainwater infiltration.
- Optimization of wastewater-based surveillance for improved public health monitoring.

## 1. Introduction

The COVID-19 pandemic has highlighted the need for effective real-time monitoring and prediction of infectious disease dynamics to support timely and informed intervention policies. In this context, various studies have been conducted to assess vaccine distribution strategies [6, 12] or the effectiveness of non-pharmaceutical interventions like telework suggestions, prohibition of private gatherings of certain sizes, or partial lock-downs [39, 7, 27, 19].

The spread of infectious diseases is nowadays modeled using a broad range of approaches, including statistical and machine learning models [39, 27, 33], compartmental and meta-population models [10, 6, 45], and agent-based models [7, 29, 22, 15], or even hybrid approaches [18, 4]. Among these approaches, agent-based models (ABMs) allow for the most detailed description of disease dynamics. ABMs simulate the spread of infectious diseases on an individual level, thereby facilitating the incorporation of comprehensive information about localization, interaction, and behavior. The models are intrinsically stochastic and based on discrete-or continuous-time Markov processes. While ABMs are the state-of-the-art in infectious disease modeling, their advancement remains an active field of research. A key challenge is the choice of model parameters.

The predictive power of models for the spread of infectious diseases depends on the available data and the ability to incorporate them into models. The number of confirmed infections is the most common data source. However, confirmed infections are affected by reporting delays [26] and subject to under-reporting [28], limiting the reliability of the resulting models [35]. While single antigen and antibody tests can be cheap, the use of these tests to study large, representative population cohorts is resource intensive. Furthermore, even the cohort might be subject to sampling bias [30, 17]. Wastewater-based surveillance presents a promising solution to these issues by capturing viral concentrations from all infected individuals within a catchment area, including those who are asymptomatic or undetected by traditional testing methods. Via detection of viral RNA in sewage, outbreaks can be identified before clinical cases are reported. This capability was demonstrated during the COVID-19 pandemic, when several national wastewater-based surveillance programs were established, e.g. the AMELAG project in Germany [37]. Wastewater-based surveillance is applicable to various diseases detectable in sewage, including poliovirus, hepatitis, norovirus, and influenza [23], but also to monitoring antimicrobial resistance [9]. Yet, infectious disease monitoring based on wastewater-based surveillance data still presents several significant challenges: The choice of the sampling location can yield different viral concentrations due to the interplay of population density and sewer infrastructure. The impact of viral degradation and variations in flow-time on measurements is not well understood. The environmental conditions, e.g. rain fall, can impact measurement and the effectiveness of established normalization strategies, such as flow-based adjustments, remains unclear. Despite these challenges, integrating wastewater data with traditional infectious disease models holds promise for improving prediction accuracy and has been attempted in several studies [8, 32].

The challenges of wastewater-based surveillance data can in principle be addressed using comprehensive computational models which provide in-depth descriptions of the spread of infectious diseases as well as wastewater dynamics. Yet, to the best of our knowledge, most published studies use relatively simple approaches. For example, Wu et al. [43] calculated a rough estimate of SARS-CoV-2 prevalence upstream of a wastewater treatment facility using the normalized viral load measured in twelve wastewater samples and assumptions about the sewer system flow volume, stool sizes, and average viral concentration in stool among infected persons. The authors concluded that prevalence in their population of interest was much higher than the confirmed case count, even under conservative assumptions, but noted that their estimate was subject to considerable uncertainty, as they did not account for the timeline of viral shedding or the loss of viral copies along sewer lines, among other factors. Hart and Halden [14] used a simplified hydrodynamic model of a city sewer network to estimate SARS-CoV-2 detectability in wastewater under different temperature-driven decay scenarios. This study highlighted the importance of appropriately accounting for viral decay when analyzing wastewater data, but the authors assumed exclusively dry weather conditions and their consideration of the relationship between SARS-CoV-2 prevalence and viral load entering the sewer system was limited; accounting for variations in viral shedding across individuals and over time was out of the study’s scope. Peccia et al. [34] compared wastewater-based surveillance data to positive COVID-19 tests and hospital admissions using a basic distributed lag time series model and found that the former led the latter data by several days. This approach highlighted the potential of wastewater data to provide early warnings of outbreaks, but relied on the assumption that the observed wastewater measurements were unbiased. Finally, Nourbakhsh et al. [32] coupled an SEIR-type compartment model of SARS-CoV-2 transmission with a simple advection-dispersion-decay model of virus concentration dynamics in a sewer system to estimate cumulative incidence in several cities based on empirical wastewater measurements and reported case data. However, because of a limited sewer system model and the use of ordinary differential equations, the model was not well adapted for scenarios such as small communities or low-prevalence settings. To the best of our knowledge, no existing study on wastewater-based infectious disease monitoring uses advance methods from the well-established field of sewage network modeling [38]. This is problematic as wastewater dynamics in sewage networks are highly complex and require advanced modeling tools for the prediction of pollutant load [2]. Accordingly, advances beyond these existing methodological frameworks could significantly enhance our understanding of disease spread and lead to more effective public health interventions.

Our work contributes to the field by integrating an agent-based model (ABM) for infection dynamics, a viral shedding model, and a detailed hydrodynamic model of sewage flow and viral load. We provide the mathematical details and a numerical implementation. Through simulation studies for a respiratory virus (Section 3.1), we investigate the effects of measurement protocols (Section 3.3 and Section 3.4), precipitation events (Section 3.5), viral decay (Section 3.6), and normalization strategies (Section 3.7) on the relationship between infection dynamics and wastewater measurements. This controlled setting enables an indepth model-based analysis and practical recommendations for real-world wastewater-based surveillance. Our findings advance the development of detailed integrative models informed by data, enhancing the accuracy and reliability of infectious disease monitoring and prediction. Furthermore, the integrated model provides a basis for coherent data integration.

### 2. Mathematical Model

To study wastewater-based surveillance data, we combine state-of-the-art models for the spread of infectious diseases and wastewater dynamics (Fig. 1). The link is established using a viral shedding model. In this section, we outline the mathematical formulations of the individual models’ components and their simulation algorithms as well as their integration. All models are dynamic and are executed on the same time axis such that a coupling with comparisons of outputs is possible.

**Figure 1:**
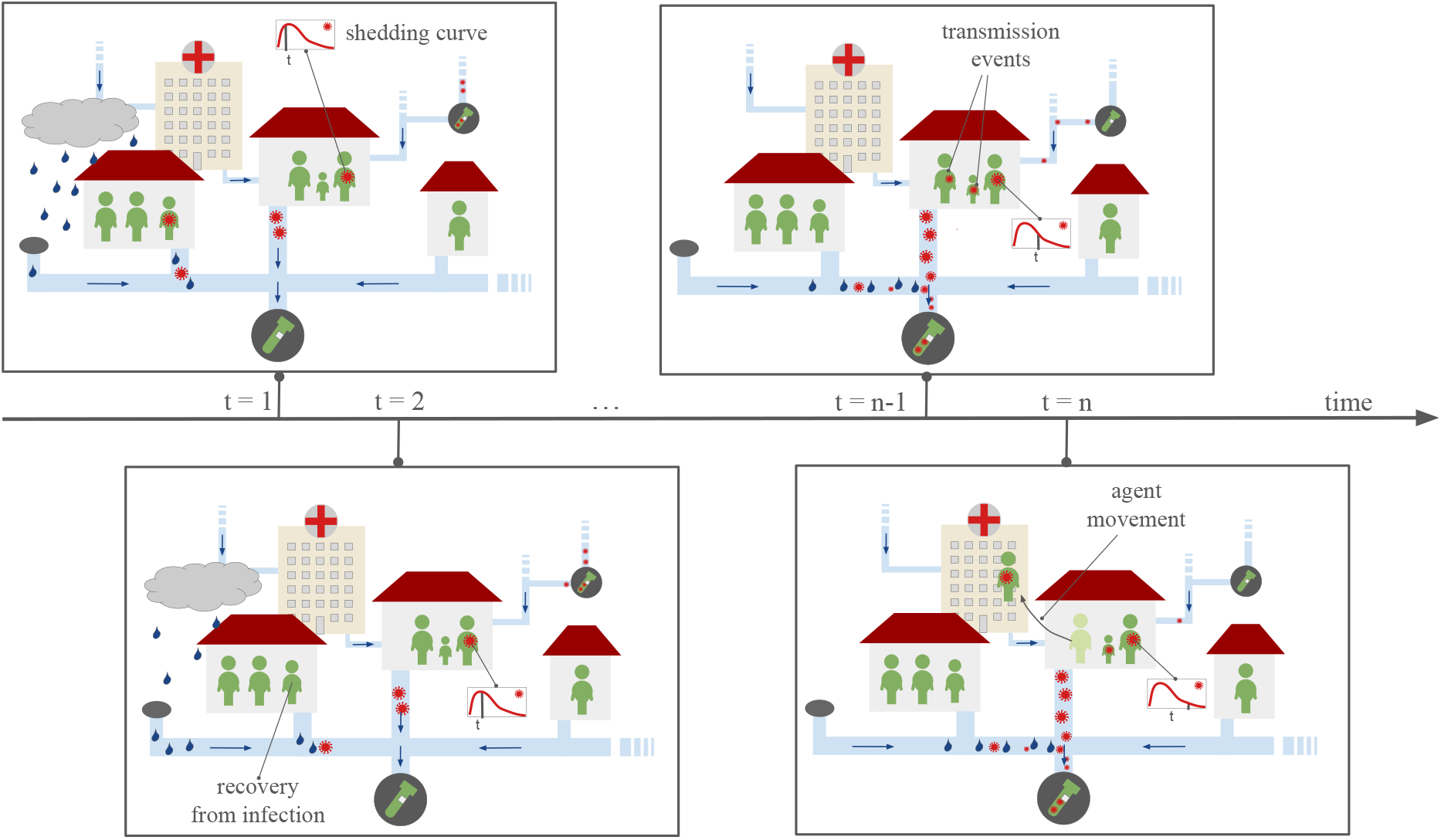
Model Visualization. Based on infection dynamics and the movement of agents, RNA is shed at specific locations and time points into the sewer system. This information is propagated over time to simulate the concentration measurements in wastewater. Among other elements, the model can account for precipitation events, sampling protocols, viral load dynamics, and RNA degradation. The figure only depicts a simplified neighborhood with locations of type home or hospital, while the model used for this study allows for various types of agent movements (Section 2.2.1).

### 2.1. Overview

The proposed model consists of three modules:

The **infection dynamics model** describes the time-dependent location and infection state of individual persons, in the following also denoted as agents. The infection state of agents can change due to events such as virus transmission, worsening of symptoms, or recovery. The likelihood of an agent infecting others is determined by its viral load, which varies throughout its infection course, and the length of contact.

The **shedding model** describes the release of virus and viral fragments from infected individuals into their surroundings. It is used in the infection dynamics model to determine the transmission probability as well as to describe the release through urine and stool. The shedding curve is assumed to be dependent on the individual’s viral load, which is time-dependent and initially increases before declining as the host’s immune response takes effect. The shedding model uses the infection state of agents and their time since transmission, which are provided by the infection dynamics model, to determine the viral RNA entering the wastewater system.

The **wastewater dynamics model** simulates the transport and degradation of viral RNA within the sewage network. Using the viral shedding input from the shedding model as well as the agents’ locations from the infection dynamics model, this module calculates the RNA concentrations at various points in the network, accounting for factors such as viral decay, flow rates, and the architecture of the sewer system. The output of this model is the RNA concentration at different sampling points, which provides a comprehensive picture of the data to be expected from wastewater-based surveillance.

In the current model, the wastewater does not influence the infection dynamics; accordingly, the integrated model possesses a hierarchical structure. In the following, we discuss the individual modules in more detail.

### 2.2. Modeling Infection Dynamics

In this study, we use an *agent-based model* (ABM) implemented in the software framework MEmilio [24] to simulate disease states and mobility patterns at the *agent* level, providing a fine-grained view of disease dynamics. It comprises agents with different attributes.

The properties of an *agent α* are defined via an *m*-tuple (*a*_1_, …, *a*_*m*_) ∈ Ω with *m* different attributes *a*_*i*_, *i* = 1, …, *m*. The attributes can be static – meaning that they do not change over the course of the simulation – or dynamic. The static attributes are:

- An agent’s age group 𝒜 ^(*α*)^ ∈ *{*1, …, *n*_*A*_*}*, with *n*_*A*_ denoting the total number of age groups.
- An agent’s set of locations 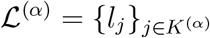 with *K*^(*α*)^ ⊂ *{*1, …, *n*_*L*_*}* denoting the subset of all locations it can theoretically move to during a simulation.

The dynamic attributes are:

- An agent’s current location *l*^(*α*)^ ∈ *ℒ* ^(*α*)^.
- An agent’s current infection state *s*^(*α*)^ ∈ *𝒮*, with 𝒮 denoting a set of infection states.
- An agent’s time since virus exposure *τ* ^(*α*)^ in hours, which is set to NaN if the agent has not been infected.

The simulation of the ABM provides information about the agent’s trajectory in space and infection state. In the following, we provide additional details on the ABM, providing the basis for the simulation of mobility (Algorithm 1) and the full population dynamics (Algorithm 2).

#### 2.2.1. Mobility Model

The ABM uses a location graph with *n*_*L*_ locations, 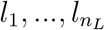, to model mobility. Every location has a *location type T* ∈ *𝒯* with 𝒯 being a set of location types such as *Home, School, Work, Recreation, Shop, Hospital*, and *Intensive Care Unit (ICU)*. There can be multiple locations of the same type. In addition to the type, a location also has a capacity specifying the maximum number of agents that can enter the location and a maximum number of contacts an agent can have at the location. Every agent *α* has a set of *n*_*α*_ many locations 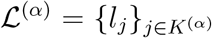, with *K*^(*α*)^ ⊂ *{*1, …, *n*_*L*_*}* being an index set, that are assigned to it. These assigned locations are the ones the agent can move between, meaning that it is restricted to a subgraph of the global location graph (see Supplementary Fig. B.1(b)). We denote the current location of an agent *α* at a given time point 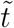, with 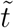 given in hours, as 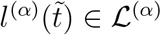. Movements, i.e. location transitions, are modeled by an ordered set of mobility rules 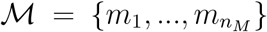 that have probabilistic components, which cause stochasticity between simulations. These mobility rules include daily regular behavior like going to work or school on weekdays, irregular behavior like occasionally attending a social event, and behavior related to the infection state of an agent, e.g. going to hospital when having severe symptoms. For Ω containing all potential states of any agent and 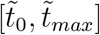 denoting the simulation period, a mobility rule 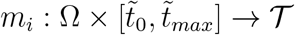 is given by

#### Algorithm 1

ABM Mobility.

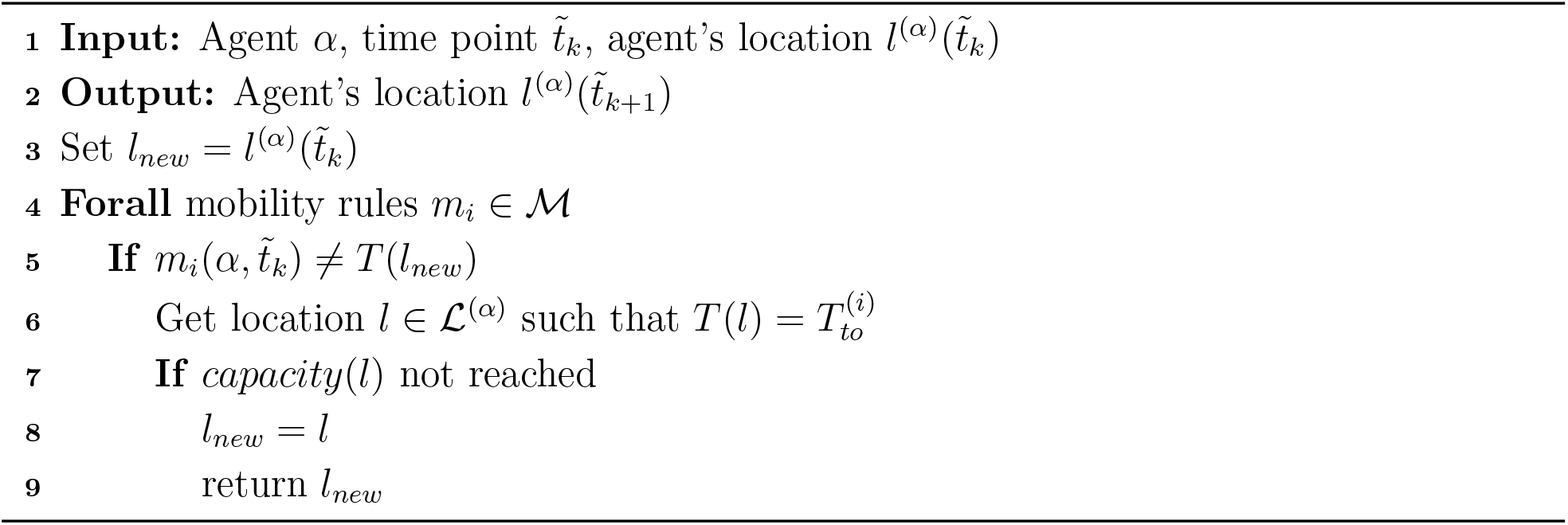

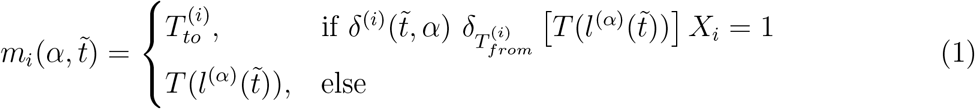

with *T* (*l*) ∈ *𝒯* denoting the type of location *l, X*_*i*_ ∈ *{*0, 1*}* denoting a Bernoulli distributed random variable with probability *p*_*i*_, which has a different value for every mobility rule, and *δ*_*_[*·*] denoting binary-valued functions defined as

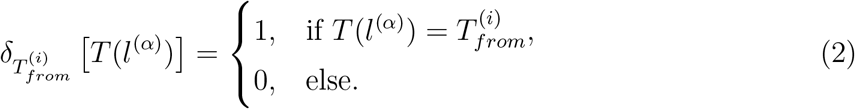

Furthermore, 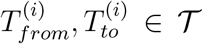 are location types and 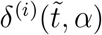 are binary-valued functions provided in Appendix A together with *p*_*i*_ for each mobility rule. The location *l*^(*α*)^ of agent *α* at time point 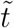, with 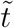in hours, is given by 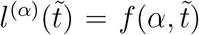 (see Algorithm 1), which is evaluated at discrete time points 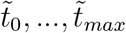 given a time step 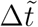 and 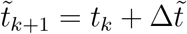.

#### 2.2.2. Disease Progression

An agent *α* has a time-dependent infection state *s*^(*α*)^(*t*) from a set of infection states 𝒮. The infection states used for this study are *Susceptible* (*S*), *Exposed* (*E*), *Non-symptomatically Infected* (*I*_*ns*_), *Symptomatically Infected* (*I*_*sy*_), *Severely Infected* (*I*_*sev*_), *Critically Infected* (*I*_*cri*_), *Recovered* (*R*), and *Dead* (*D*). Infection state *I*_*ns*_ includes infectious pre- and asymptomatic agents and infection state *I*_*sev*_ includes agents requiring hospital treatment while *I*_*cri*_ includes agents requiring ICU treatment. We call an agent *infected* if it has infection state *s*^(*α*)^(*t*) ∈ *{E, I*_*ns*_, *I*_*sy*_, *I*_*sev*_, *I*_*cri*_*}* and *formerly infected* if *s*^(*α*)^(*t*) ∈ *{R, D}*. Transitions between infection states are stochastic and possible either through virus transmission (*S* → *E*) (see Section 2.2.3) or disease progression (*E* → *I*_*_, *I*_*_ → *I*_**_, *I*_**_ → *{R, D}*) (see Supplementary Fig. B.1(a)). For a (formerly) infected agent *α*, the course of infection is defined as 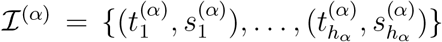 containing the time points 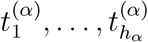 at which the agent changes or changed its infection state and the corresponding infection states 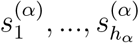. Hence, 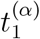 is the time point at which the agent is exposed and 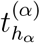 the time point at which the agent recovers or dies, i.e. 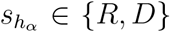. The intermediate time values 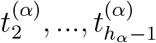 are the time points at which an agent changes to one of the infectious states of the agent’s individual course. The length *h*_*α*_ of the course of infection differs between non-symptomatic and (severe or critical) symptomatic courses and is therefore agent-dependent. The stay times in infection states *E*, …, *I*_*cri*_ are log-normally distributed and the transitions *E* → *I*_*_, *I*_*_ → *I*_**_, *I*_**_ → *{R, D}* between all infection states, apart from virus transmission (*S* → *E*), are Bernoulli distributed; see Supplementary Table C.1 for the values of all disease progression-related parameters used in the results section.

#### 2.2.3. Disease Transmission

Infected agents can transmit the virus to susceptible agents if they are at the same location. For a susceptible agent in location *l*, the waiting time until transmission is exponentially distributed with rate Λ_*l*_(*t*). Assuming that agents change their locations only at discrete time points, *t*_0_, …, *t*_*max*_, the number of agents at a location is constant in [*t*_*k*_; *t*_*k*+1_) and Λ_*l*_(*t*) is given by

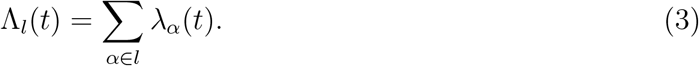

for *t* ∈ [*t*_*k*_; *t*_*k*+1_) given in days. We use the casual notation *α* ∈ *l* to iterate over the infected agents at location *l* in the interval [*t*_*k*_; *t*_*k*+1_). The agent-dependent rate *λ*_*α*_(*t*) is given by the infectiousness curve described in Section 2.3, Eq. (8). The location-specific infection rate (3) builds on the assumption of homogeneous mixing within the location. If the waiting time until transmission is longer than the time until the susceptible agent leaves location *l*, no transmission occurs. Hence, for a fixed time step Δ*t* = *t*_*k*+1_ − *t*_*k*_ given in days, the probability that a susceptible agent at location *l* gets exposed is

##### Algorithm 2

BABM Simulation.

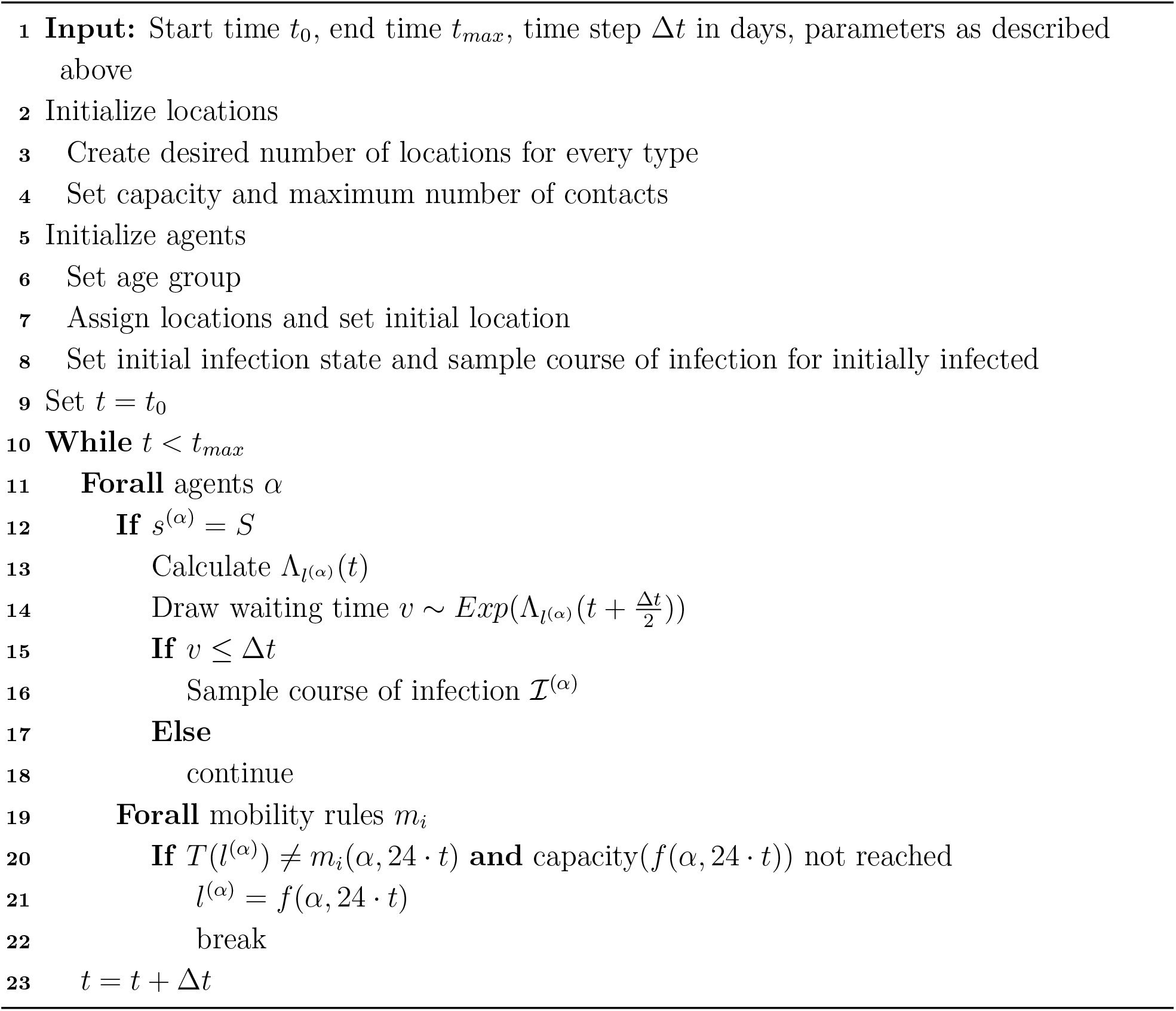

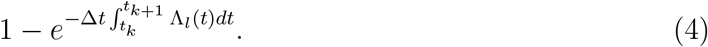

### 2.3. Modeling Viral Load, Infectiousness and Shedding

The viral load of individual patients determines their infectiousness and shedding. Here, we model viral shedding using established models [20, 21]. The viral load for an agent *α* in RNA copies per swab on the *log*_10_ scale at a given time *t* in days (see Supplementary Fig. B.2, left) is defined as

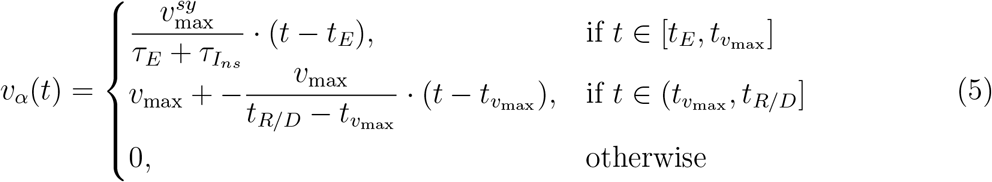

with 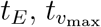, and *t*_*R/D*_ denoting the times in days of virus exposure, maximal viral load, and recovery/death, respectively. The time points are agent specific, with 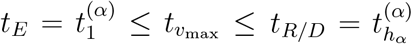. As the infection state trajectories are sampled at the time point of exposure, *t*_*R/D*_ is readily available.

We assume that symptomatically infected agents reach the peak viral load the moment they show symptoms, while agents not showing symptoms reach the peak in the middle of their infection period. Furthermore, we assume agents that share the same duration 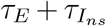 also share the same linear increase until 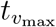. This yields for the variables 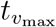 and *v*_max_:

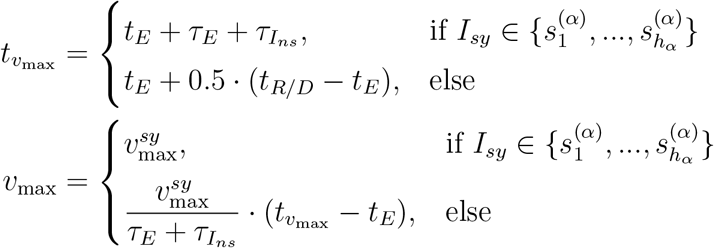

where 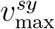 is the peak viral load given in *log*_10_ RNA copies per swab for symptomatic infections (see Supplementary Table C.2 for the value of this and all other parameters relevant to viral shedding).

Following the observation of Jones et al. [20], we model the shape of an agent *α*’s shedding by a sigmoid function of their viral load, i.e.,

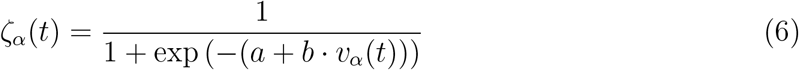

with shape parameters *a, b >* 0. As in [20], since there is no information about when shedding starts after exposure, we make the assumption that shedding is zero or close to it as long as the agent is still in the Exposed state. Therefore we introduce a time shift of *τ*_*shift*_ = 0.6 *· τ*_*E*_. Thus, the scaled and shifted shedding curve for an agent *α* is given by:

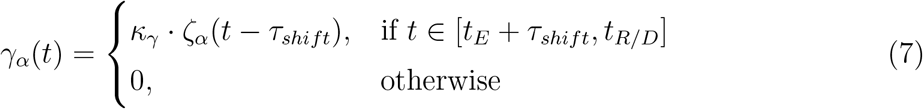

where *κ*_*γ*_ is a scaling factor that translates *ζ*_*α*_(*t* − *τ*_*shift*_) into an RNA shedding rate. The RNA shedding into the sewage network is the number of RNA copies shed in total per day and has to be normalized by the water flushed into the wastewater system to receive a unit of copies per liter.

Similarly, the corresponding unitless infectiousness curve for agent *α* at time point *t* (given in days) (see Supplementary Fig. B.2, right) is given by

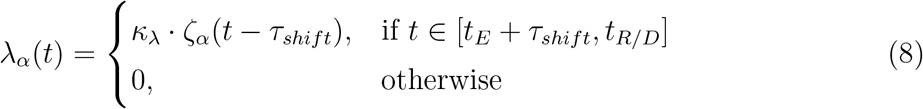

with *κ*_*λ*_ translating *ζ*_*α*_(*t* − *τ*_*shift*_) into a transmission rate (see Section 2.2.3).

### 2.4. Modeling the Sewage System and its Hydrodynamics

We use a comprehensive model of a wastewater network to simulate the sewage flow and reactive transport of dissolved chemical substances in wastewater while avoiding simplifications and incorrect interpretations. In this model, a sewer system is represented as a directed acyclic graph defined by *n* edges ℰ = *{e*_1_, …, *e*_*n*_*}* and *m* nodes 𝒩 = *{n*_1_, …, *n*_*m*_*}*. The edges represent sewage pipes and the nodes represent junctions as well as entry and exit points. Edges and nodes are characterized by several parameters, including total volume and height. The state of the system is the amount of water contained in the edges and nodes, its flow rate, and the concentrations of the relevant substances. The simulation of water inflow to the system is based on two phenomena: hydraulic surface runoff during precipitation events and water usage of industry and citizens.

At their core, the hydrodynamic calculations are based on the Saint-Venant Equation [16]. The equation assumes one-dimensional flows in open channels and mass and momentum conservation, which yields

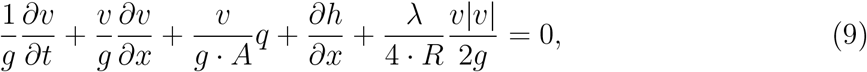

with flow velocity *v* in 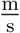, time *t* in s, gravitational acceleration *g* in 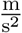, vertical position along the pipe *x* in m, cross-sectional flow area *A* in m^2^, lateral inflow corresponding to the precipitation of a specified time interval *q* in 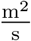, water levels *h* in m, pipe friction coefficient *λ* (unitless) and hydraulic radius *R* in m (i.e. 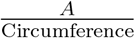).

Replacing the differential operators with differential quotients for specified time points *t*_1_ *< t*_2_ and locations *x*_1_ *< x*_2_ yields

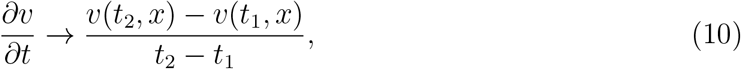

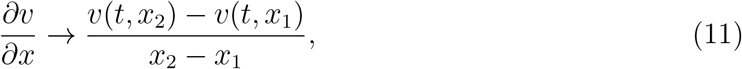

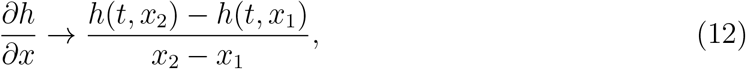

for *t* ∈ [*t*_1_, *t*_2_] and *x* ∈ [*x*_1_, *x*_2_]. By interpreting Δ*x* = *x*_2_ − *x*_1_ as the length of an edge of the wastewater network, Eq. (9) after rearrangement becomes a quadratic equation of the form

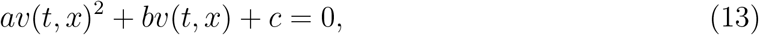

where

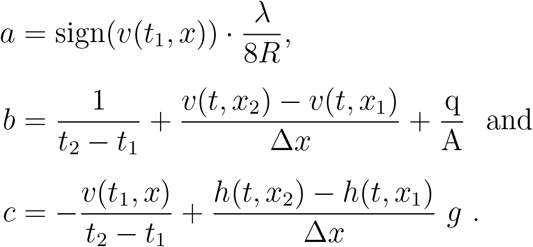

The quadratic equation has two complex solutions, where the flow velocity *v* is equal to the real part of those solutions:

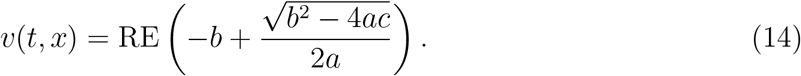

Using this solution and the boundary conditions *v*(*t, x*_1_), *v*(*t, x*_2_), *h*(*t, x*_1_), and *h*(*t, x*_2_), as well as the initial condition *v*(*t*_1_, *x*), a solution of the flow velocity can be calculated for arbitrary time points and edges of the network system. The state of the nodes of the system define the boundary and initial conditions. Based on the solutions from Eq. (9) for edges connected to a node, the in- and outflow to the node is given for each edge. At the center of the node, the sum of in- and outflow equals zero (mass conservation). Based on this assumption, the flow rates and heights at the edge borders can be calculated.

While Δ*x* can be defined based on the length of an edge, Δ*t* = *t*_2_−*t*_1_ has to be chosen carefully. A too large value of Δ*t* will yield inaccurate numerical solutions, while a too small value will yield unnecessarily high computation times. A complex solution of Eq. (13) with an imaginary component larger than zero indicates the transition to an oscillatory state. To ensure a meaningful numerical solution, Δ*t* is chosen to be smaller than the corresponding oscillation period. Further considerations like the maximum total change of volume provide an equation to set Δ*t* for each iteration of the numerical solution scheme, such that stable solutions of *v* are ensured even for extreme hydraulic scenarios (see [41]). By combining Eq. (13) with other known physical principles, e.g. energy loss along a pipe (using the Prandtl-Colebrook equation) and energy loss due to inelastic collisions in manhole structures (according to the Borda-Carnot equation), the simulation precision is further refined.

Based on the calculated flow velocities and other time-varying edge state variables, the substance concentration per location and time point can be calculated. Viral loads generated by agents of the ABM enter the sewer system as concentrations in the respective amount of domestic wastewater generated for every time step of the model simulation, at the location the agent currently occupies. Viral fragments are then transported through the system according to the pre-calculated flow rates, potentially taking chemical reactions in the form of viral decay into account. The outputs of the hydraulic simulation are time- and location-dependent concentration curves. The time step for the viral load calculation is chosen prior to the calculations and a suitable value depends on the specifics of the viral decay dynamics, where faster changing dynamics suggest choosing a smaller time step.

After defining the wastewater network, its connected surfaces and corresponding runoffs, as well as substance characteristics, the simulation proceeds in two steps (see Supplementary Fig. B.3). First, the flow velocities and volumes are calculated with a numerical solver. Secondly, the viral load over time is simulated.

For the numerical simulation, we use the urban water management modeling and simulation environment ++SYSTEMS, developed by the company tandler.com GmbH. Utilizing the mathematical principles described above, ++SYSTEMS with its backend and calculation kernel DYNA forms a fully dynamic, geospatial modeling and management software for waste- and rainwater (individually or combined) sewer systems. Details on the implementation are available in [41].

## 3. Results

### 3.1. Demonstrator Setup

To address open questions and challenges related to the interpretation of wastewater-baser surveillance data, we performed a simulation study, which allows us to evaluate counterfactual scenarios without missing data or data uncertainty. To this end, we developed a synthetic, yet realistic model of a city neighborhood and corresponding sewer system (Fig. 2); we then traced infectious disease outbreaks in this controlled setting using our sequence of three modules, from the MEmilio-based infection dynamics model to the shedding model to the ++SYSTEMS-based wastewater dynamics model. The synthetic neighborhood used in our study contains residential, recreational, university, mixed shopping and business, and mixed residential and industry surface areas. It is populated by at least 838 agents, whose simulated movements, both on weekdays and weekends, remain completely inside the model. The model’s synthetic sewer system was designed such that realistic sewer conditions are maintained during all simulation scenarios: No sanitary sewer overflow occurs, all pipes are at a maximum of 90% of their hydraulic capacity, and gravity flow is realized throughout the whole system.

**Figure 2:**
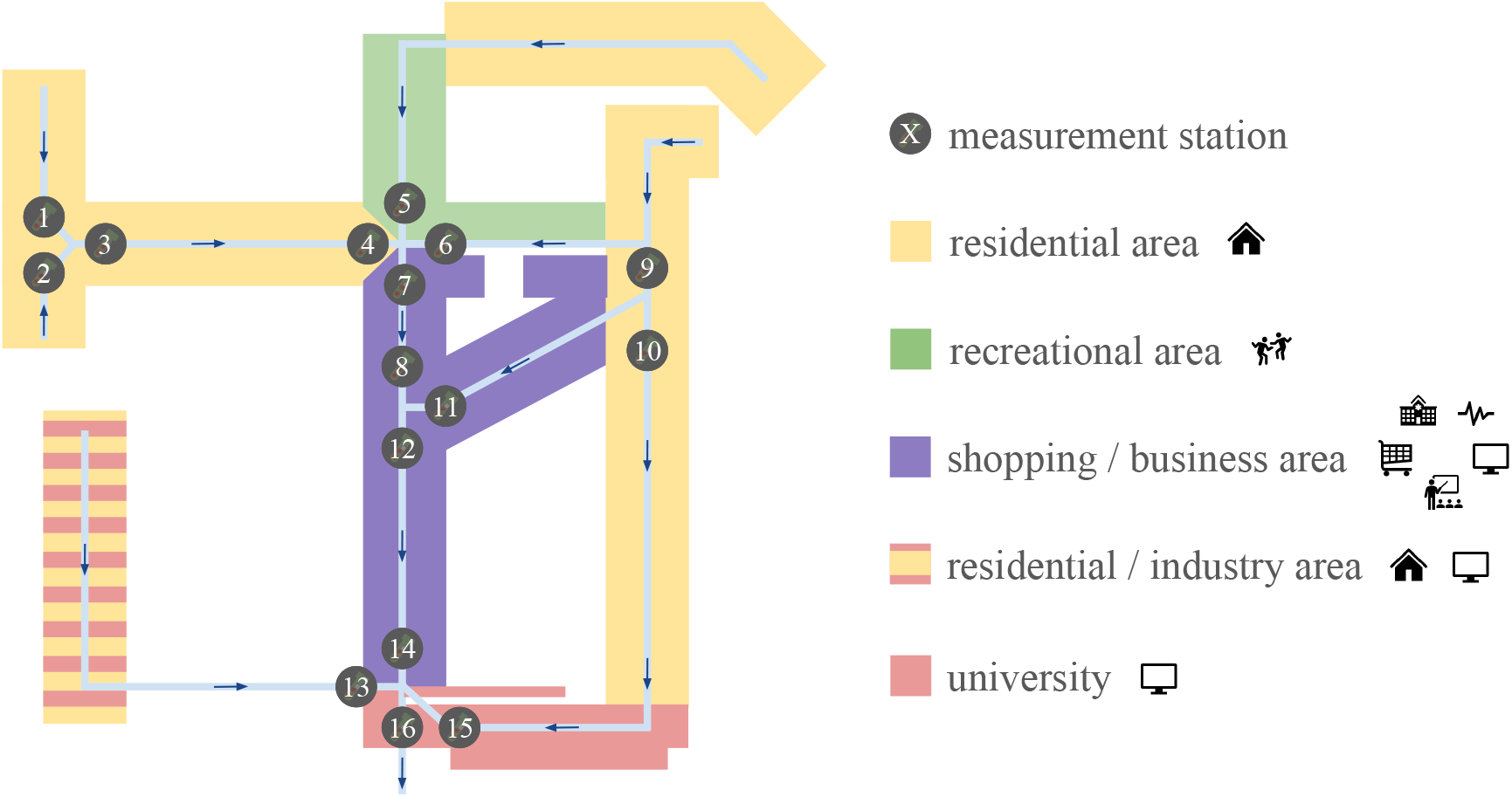
Demonstrator Neighborhood. The synthetic neighborhood on which we base our simulation study constitutes areas of different types. Flow rates and substance concentrations are simulated for 16 different measurement stations.

Following the official reporting standards for COVID-19 cases in Germany, we considered *n*_*A*_ = 6 age groups ranging from small children to seniors. Households – i.e. groups of agents that share their assigned *Home* location – were created for each residential area based on its total number of agents. We considered 1- to 5-person households. Every household had at least one member of the adult age groups 𝒜 ∈ *{*3, 4, 5, 6*}* (age groups 1 and 2 correspond to early childhood or adolescence). The household distribution was motivated by the German micro census 2019 [40]. Non-*Home* locations also had to be assigned to the agents. A location of a given type was assigned to an agent from an equal distribution of all locations of that type. Every agent was assigned a location of type *Shop, Recreation, Hospital*, and *ICU*, while a *School* location was only assigned to agents in age group 2 and a *Work* location only to agents in age groups 3 and 4. Finally, the initial infection states were allotted to agents by independent and identical sampling from the initial infection state distribution (0.2% *E*, 0.5% *I*_*ns*_, 0.29% *I*_*sy*_, and 0.01% *I*_*sev*_), i.e. on average, 1% of the modeled agents were initially infected.

The integrated model contains various parameters describing characteristics of the virus, which allows for the modeling of a broad spectrum of communicable respiratory or, in particular, COVID-19-like diseases. We implemented the simulation using wild-type COVID-19-like parameters based on [25], [22], and [21]. The ++SYSTEMS files defining the exact shape of the sewage system, area characteristics, etc. are provided as supplementary material for each experiment and in the subsequent sections we only mention settings that differ between the experiments. An overview of the ABM parameters as well as the experiment-specific parameters is provided in Appendix C.

Since most cluster systems are based on Linux, we facilitate the modules using Ubuntu. ++SYSTEMS is a Windows program, hence, we created a headless virtual machine, which can be started and navigated through via a command line interface. One ABM simulation takes about 3.8 minutes on one core; one ++SYSTEMS simulation takes about 3-4 minutes on 8 cores.

### 3.2. Non-trivial relation of prevalence in catchment area and measurement concentrations

For a first assessment of the process dynamics, we considered the total number of upstream agents, the number of upstream infected agents, and the upstream RNA influx for a single measurement station for an outbreak scenario (Fig. 3). The model simulation reveals that the number of agents in a particular catchment area changes over the course of a day and from weekdays to weekends, primarily due to the agents’ participation at work, school, or recreational events. This mobility results in substantial changes to the upstream RNA influx. The measured virus levels are additionally influenced by the sewage volume, which itself depends on the total number of agents in the catchment area. Since agents return home from school, etc. at slightly different time points, the virus levels can show large deviations from an average value for only a few simulation minutes. Overall, the model highlights the impact of mobility on wastewater-based surveillance results.

**Figure 3:**
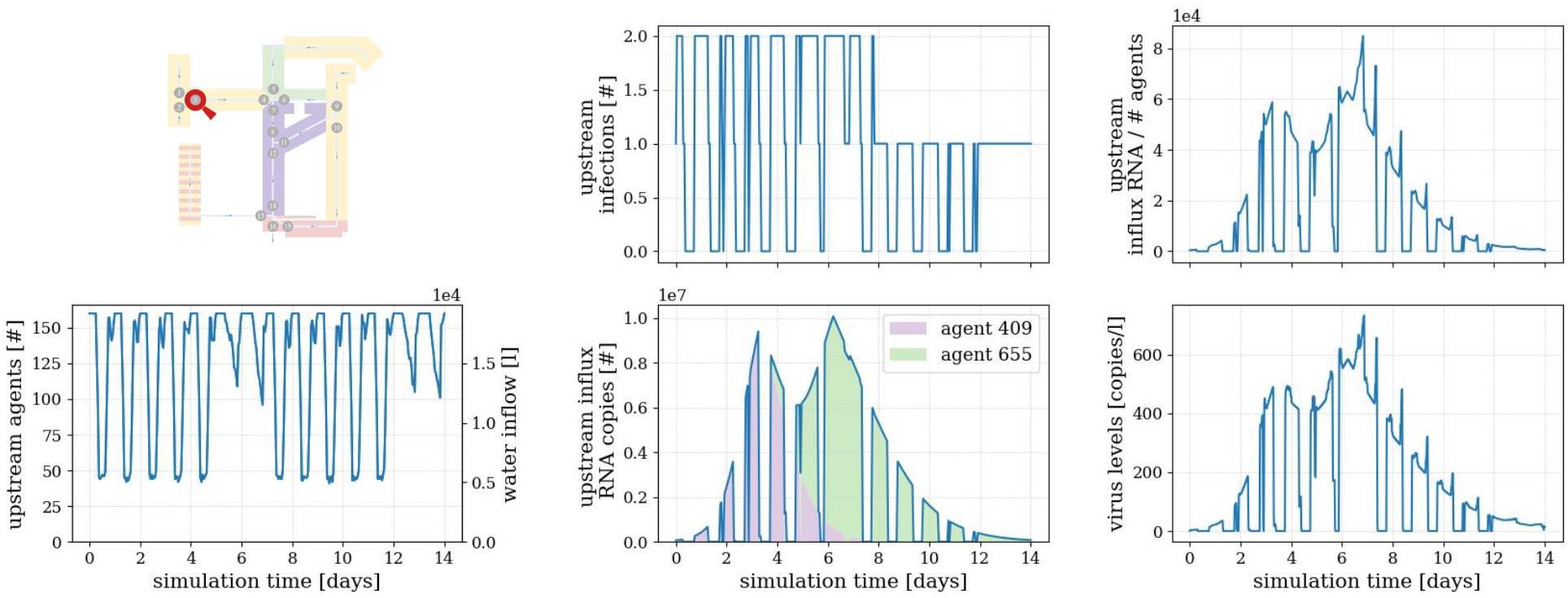
Exemplary (processed) Simulation Output. Output for viral measurements in a scenario without precipitation or viral decay at measurement station 3.

### 3.3. Characteristics of catchment area influence dynamics

Establishing a monitoring system for wastewater is time- and energy-consuming. Legal permits for measurements and access to the locations have to be organized, the sampling stations have to be set up, and the samples have to be collected and transported to laboratories for further analysis on a regular basis. Hence, it is not surprising that even established monitoring networks have limited sampling locations and time schedules, often reporting 1-3 values per week and neighborhood or city. This renders the optimal placement of sampling locations and the selection of appropriate sampling strategies critical. Here, we investigated the impact of the choice of sampling location using our fine-grained integrated model, as a corresponding real-world study would be infeasible.

We considered 16 possible sampling location in the synthetic neighborhood. These sampling locations correspond to catchment areas with a broad spectrum of different properties: First, the corresponding catchment areas of a sampling station differ with respect to the area type, i.e. primarily include residential areas (stations 1, 2, 3, 4, 9, 10), recreational areas (station 6), shopping/business areas (station 11), or mixed areas. Second, the catchment areas differ with respect to their sizes. Further upstream locations (e.g. station 1) summarize the dynamics of a smaller area than downstream locations (e.g. station 16). In this neighborhood, sewer flow times to the furthest downstream stations are at most around 80 minutes.

To assess the information content of the wastewater-based surveillance data, we conducted a comprehensive simulation study. A total of 250 simulations of the proposed integrated model were used to account for the inherent stochasticity of infection processes; see Supplementary Fig. B.4 for a visualization of the prevalence over time. Assessment of the simulation results (Fig. 4) shows that sampling locations downstream of residential areas (e.g. station 1) produce reproducible daily and weekly trends in the measured virus levels. In contrast, sampling locations downstream of regions containing recreational areas (e.g. station 6 & 11) show more variability between simulations. Sampling stations near the endpoint of the network, which have large catchment areas (e.g. station 16) and would in practice fall closer to a wastewater treatment plant, yield smoother curves with no or less extreme daily and weekly trends.

**Figure 4:**
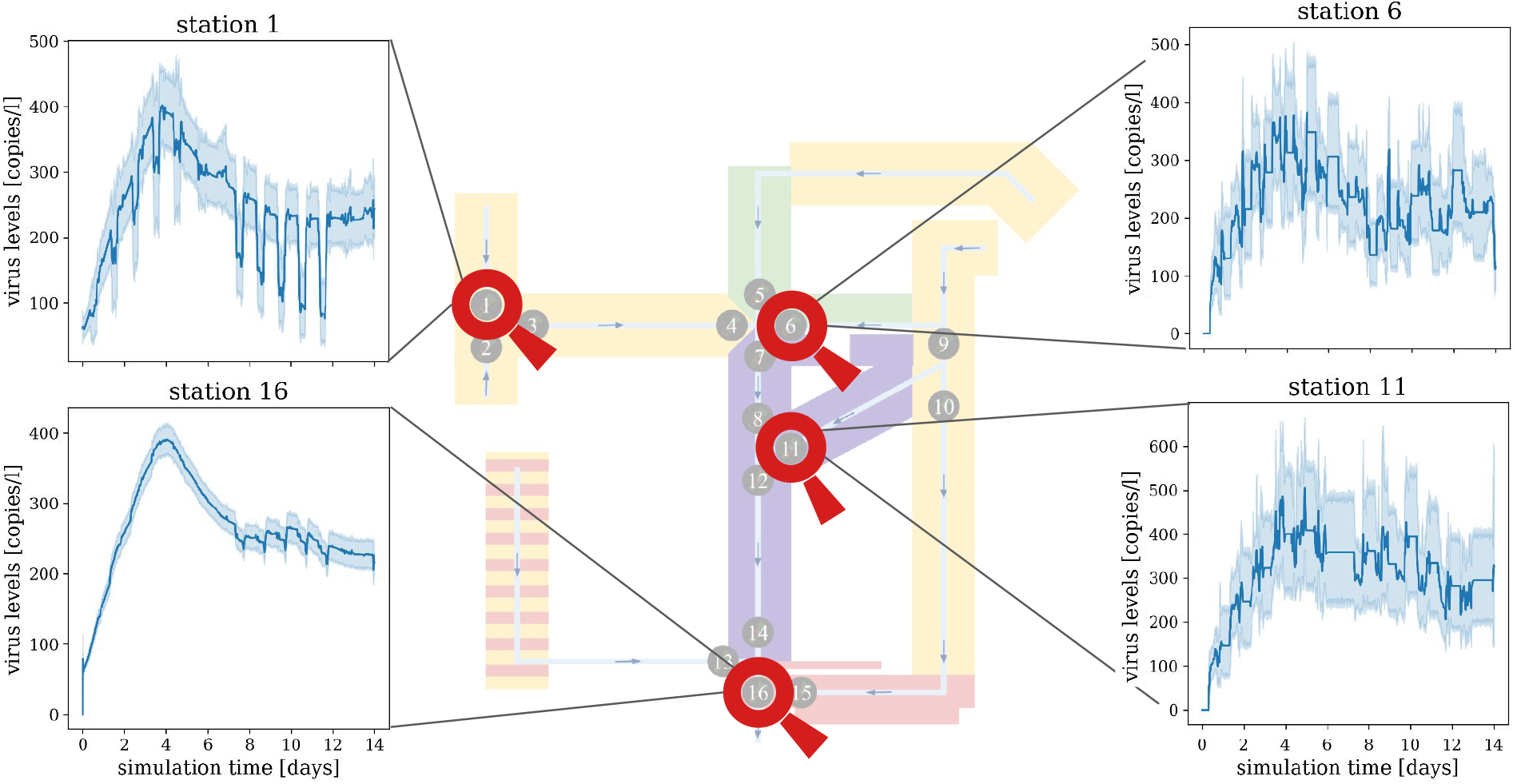
Variability of Measurements for Sampling Locations. Comparison of virus levels in wastewater at different sampling locations. Shown are the mean (solid line) and 95% confidence intervals (shaded area) of the 250 simulation results per location.

To evaluate how representative the viral load in the wastewater is at the different sampling locations, we computed the temporal cross-correlations between the RNA copies per liter in wastewater samples and (i) the true overall prevalence (Fig. 5(a), top) and (ii) the true viral shedding into the wastewater (Fig. 5(a), bottom). We found the highest cross-correlation values are reached for the stations with larger catchment areas, in particular stations 7, 8, 12, 14, and 16. The correlation coefficient is as high as 0.56 for the true prevalence and 0.90 for the amount of virus shed. These corelation coefficients are surprisingly high given the variability between the (stochastic) simulation runs (Fig. 5(b)). Indeed, if the initial infections were not distributed randomly, the pattern would be more pronounced and large catchment areas would be even more beneficial (results not shown).

**Figure 5:**
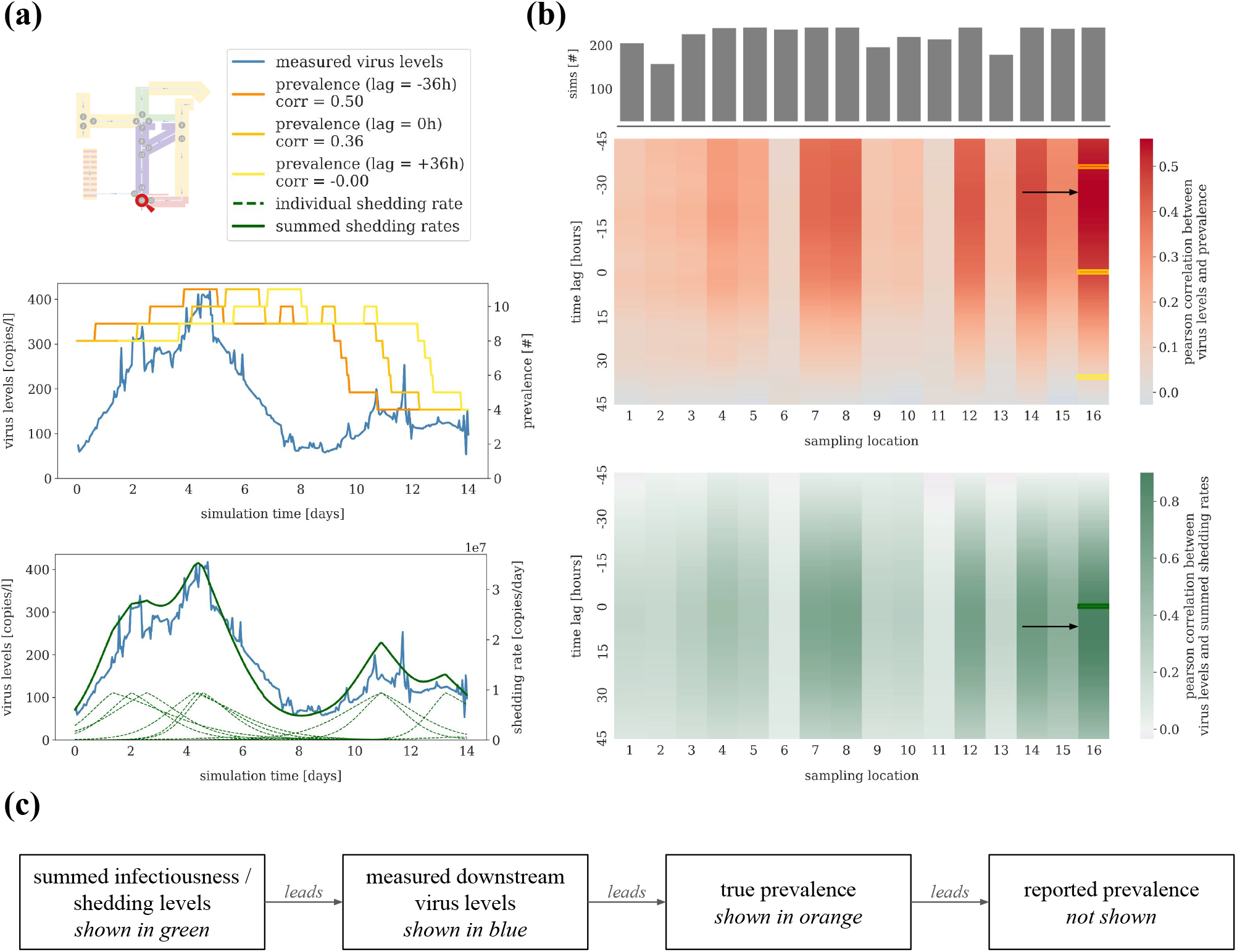
Cross-Correlations Between Wastewater-based Surveillance Data and Prevalence. (a) Trajectory of the wastewater viral load in RNA copies per liter for sampling location 16 compared to the total true prevalence shifted with lags -36, 0, and 36 hours (top), or to the summed shedding rates across all prevalent infections (bottom), for one simulation. (b) Pearson cross-correlations between RNA copies per liter in wastewater measured at the 16 different locations and the total true prevalence (top) or the summed shedding rates (bottom), averaged over 250 simulations. The time lag describes the shift in prevalence or shedding rates. The maximum cross-correlations are marked with black arrows. The upper bar plot indicates how many simulations were used to calculate the correlations for each sampling location; simulations in which no virus was ever measured at a particular location were removed. (c) Schematic illustrating the temporal relationship between the different outcomes.

The integrated model also shows that the temporal cross-correlation is generally higher when a negative time lag is applied to the (true) prevalence data. Since a high virus concentration in the wastewater indicates that the level of infectiousness across the population is also high and that a wave of new infections will therefore likely soon follow, the virus level at sampling stations with large catchment areas was most predictive for the prevalence 10 to 40 hours later. This time lag likely depends on the incubation time of the virus and its replication rate in the human body. As the reported prevalence is delayed compared to the true one, the time shift observed in practice will be even larger (Fig. 5(c)). Overall, our results suggest that in the absence of complicating factors such as viral decay – the impact of which would be limited in this particular sewer due to the relatively short travel times – choosing a wastewater sampling location far enough downstream to be unaffected by daily and weekly trends may help predict increases in prevalence before they occur.

### 3.4. Temporal sampling design has minimal impact on wastewater monitoring results if samples are taken downstream

The sampling design differs between wastewater monitoring studies. The most common setups are one grab sample collected per day of interest (usually during the morning flush) or a 24-hour compound sample based on a collection of one sample per hour [23]. To assess the impact of these sampling strategies and their benefits and disadvantages, we simulated both strategies using the same setup as in the previous section (in which we assumed the use of discrete grab samples every three minutes). The analysis of the simulation results indicates that in the case of a clear daily trend, the choice of sampling protocol can influence the results and e.g. lead to systematically biased estimates of the general dynamics (Fig. 6). Stations further downstream are less influenced by daily and weekly agent movement patterns (see Section 3.3) and different sampling protocols yield comparable results, i.e. a maximal cross-correlation of 0.56 between measured viral load and time-lagged prevalence.

**Figure 6:**
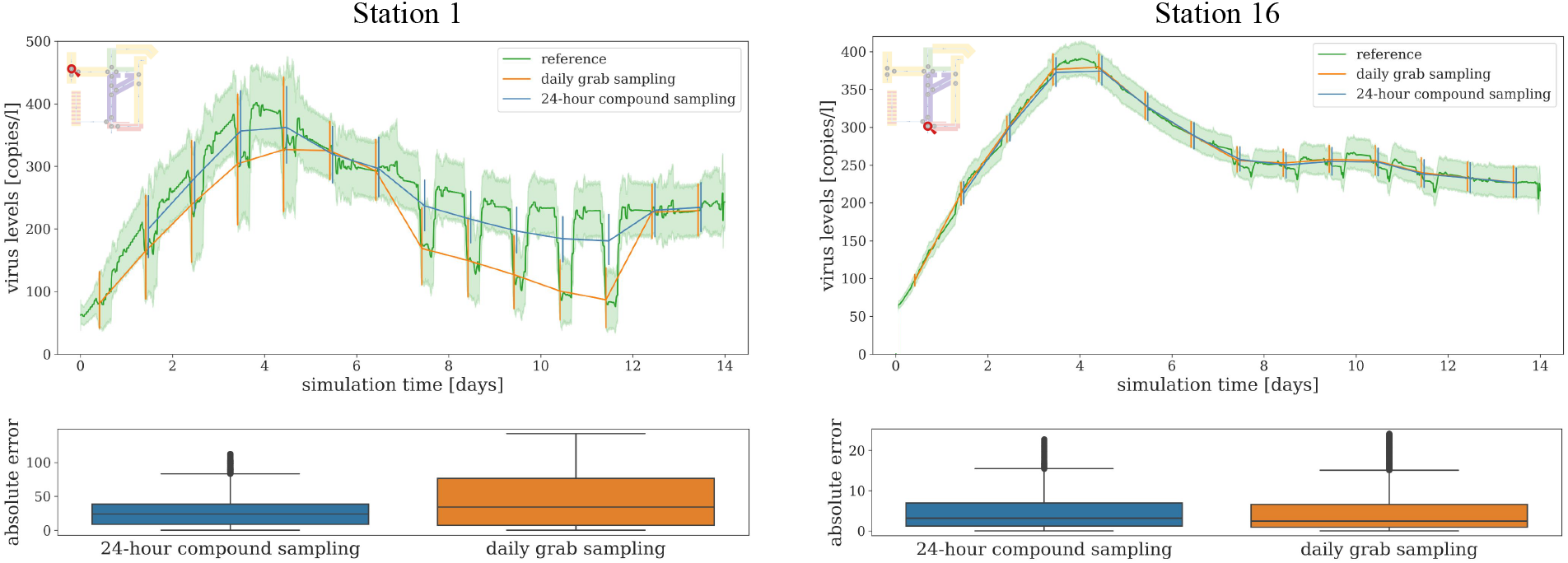
Sampling Protocols. RNA concentration in wastewater with 24-hour compound sampling or daily grab sampling (at 10:00 am each day) compared to the reference scenario (grab sampling every three minutes). Shown are the mean (solid lines) and 95% confidence intervals (shaded areas or error bars) of the 250 simulation results for stations 1 and 16 along with the distribution of the absolute error between the linearly interpolated average results for 24-hour compound or daily grab sampling and the average results for the reference scenario.

### 3.5. Rain influx impacts reliability of wastewater monitoring results in a nonlinear manner

Rain influences the amount of fluid in the wastewater system, the fluid velocity, and the concentration of particles in the overall wastewater. Yet, many currently available models simply disregard rain events and the associated dataset. This approach results in a loss of information, and – as it is unclear how long-lasting the effects of rain might be – might still not be particularly reliable. To provide a fine-grained analysis of the impact of rain on wastewater measurements, we simulated three scenarios: *No precipitation* (which was also used for the previous results), *moderate gentle rain*, and *moderate rain* showers. Here, we followed the rain intensity definitions from Germany’s National Meteorological Service (DWD) [42]: “moderate gentle rain” means between 0.1 mm and 0.5 mm in 60 min and “moderate rain” means between 2.5 mm and 10.0 mm in 60 min.

In order to run simulations with a two-week duration with realistic time-dependent rain intensities, a suitable two-week period from a synthetic rain series [3] generated by the Bavarian Environment Agency (LfU) was used. The intensities were adapted such that the abovementioned rain definition criteria were met. As a result, the two rain scenarios only differ in their intensities; the temporal profile of rain peaks is the same for both scenarios, ensuring comparability.

Due to effects like evaporation, the filling of water basins (e.g. uptake by the ground), and permeable and non-permeable surface fractions, only a small proportion of rainfall ends up in the hydraulic system and a minimum amount of precipitation is necessary to have an effect at all. This net hydraulic surface runoff increases the water volume and hence, can increase the flow rates and reduce the concentration of RNA in wastewater significantly (Fig. 7). RNA copies per liter close to zero correspond to a proportion close to 1 of rainwater in the sewage. If the flow rates increase by several orders of magnitude for a short amount of time (as is, e.g. the case in the moderate gentle rain scenario presented here), the sewage containing large fractions of rainwater is flushed out of the system very quickly, yielding a state comparable to the non-precipitation state afterwards. The moderate gentle rain scenario showcases that there is a minimum amount of precipitation necessary to have an effect on the flow rates and virus concentrations. The moderate rain scenario highlights how precipitation influences measurements if the rainwater inflow to the sewage is larger than zero for several neighboring time steps.

**Figure 7:**
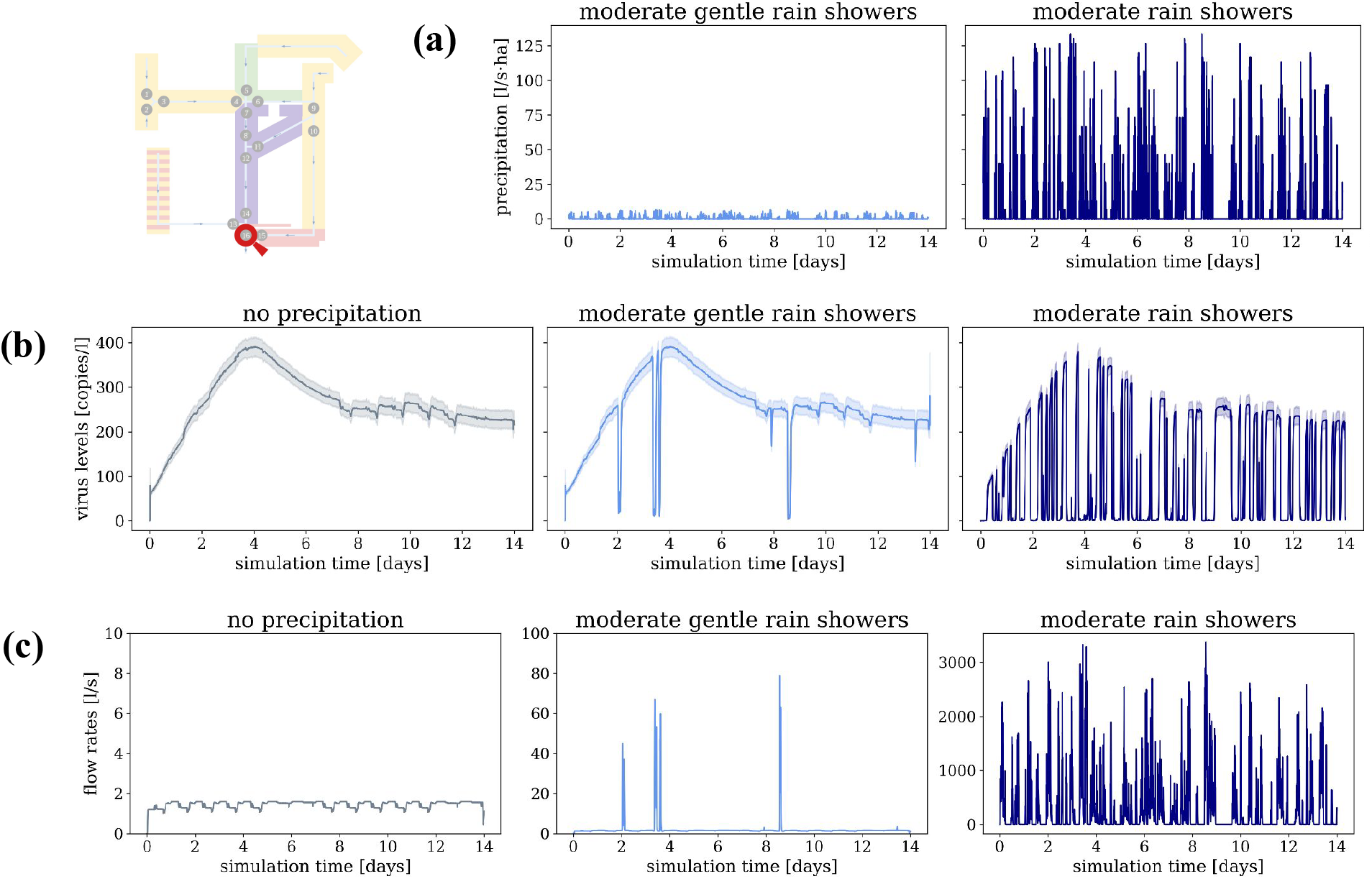
Influence of Precipitation. Comparison of different rain scenarios (no precipitation, moderate gentle rain showers, and moderate rain showers) at sampling location 16 visualizing (a) the precipitation, (b) the RNA concentration in wastewater in copies per liter, and (c) the flow rates in liter per second.

The analysis of the influence of precipitation events on measurements showcases the importance of normalizing observations to compare measurements of precipitation and dry weather time points. For more details on normalization strategies, see the results presented in Section 3.7.

### 3.6. Virus Characteristics

The interplay of virus and host immune response determine virus shedding and transmission and, hence, the prevalence and influx of virus particles into the wastewater system. Yet, the virus particles are not necessarily stable but can decay. For SARS-CoV-2, estimates of the the 90% reduction times in wastewater at ambient temperatures range between 5.5 and 28.8 days [5, 13, 1]. As the sewer of the synthetic neighborhood has a relatively short maximum flow time, we have been able to reasonably neglect viral decay so far. We now assume that our virus of interest has much faster virus reduction times than SARS-CoV-2, in order to study the impact of rapid decay processes on wastewater monitoring results and their relation to prevalence. To this end, we compare three different temporal models for the decay rate *v* of [RNA] (in copies per liter) in wastewater:

- no decay: 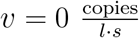
- linear decay: *v* = *k*_1_, where 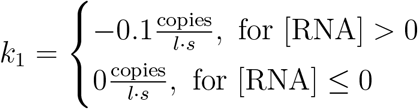
- exponential decay: *v* = *k*_2_ *·* [RNA], where 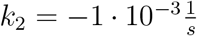 (corresponding to a half-life of about 690 seconds or about 0.2 hours)

The *no decay* scenario assumes that viral particles remain intact and serves as a baseline for understanding the upper bound and comparing it to more realistic models. The *linear decay* scenario is motivated by a potential interaction between a virus and a certain enzyme or environmental condition in the wastewater that degrades the virus at constant rate (e.g. because its abundance is limited). The *exponential decay* scenario captures the most commonly observed decay dynamics, translating to a constant decay probability per unit time.

We analyzed the simulation results for the three decay scenarios, without precipitation, at two sampling locations: 1 and 16 (Fig. 8). At the upstream station 1, the measured viral loads are proportionally slightly lower for the linear decay setting and considerably lower for the exponential decay setting compared to the no decay setting, since for the relevant concentrations, exponential decay with a half-life of 690 seconds is considerably faster than linear decay of -0.1 copies per liter per second. However, the general shape of the viral load trajectory at station 1 over time is unaffected by the decay setting: For all three decay scenarios, there are periodic dips in the virus level on weekdays and a defined peak around the fourth day. At the downstream station 16, not only are the measured viral loads lower for the linear and especially the exponential decay settings, but the shape of the viral load trajectory over time is also affected, with defined peaks during weekday daytime periods. When the viral decay is non-negligible, virus copies shed from the upstream residential areas tend to decay before they reach the furthest downstream station, so station 16 primarily measures the copies shed from the nearest areas: a university and a shopping/business region, which are only inhabited during working hours. This phenomenon indicates that the interaction between viral load and viral decay can be complex and that a detailed sewer model is necessary to adequately describe its impact on the reliability of wastewater viral loads as an indicator of disease prevalence.

**Figure 8:**
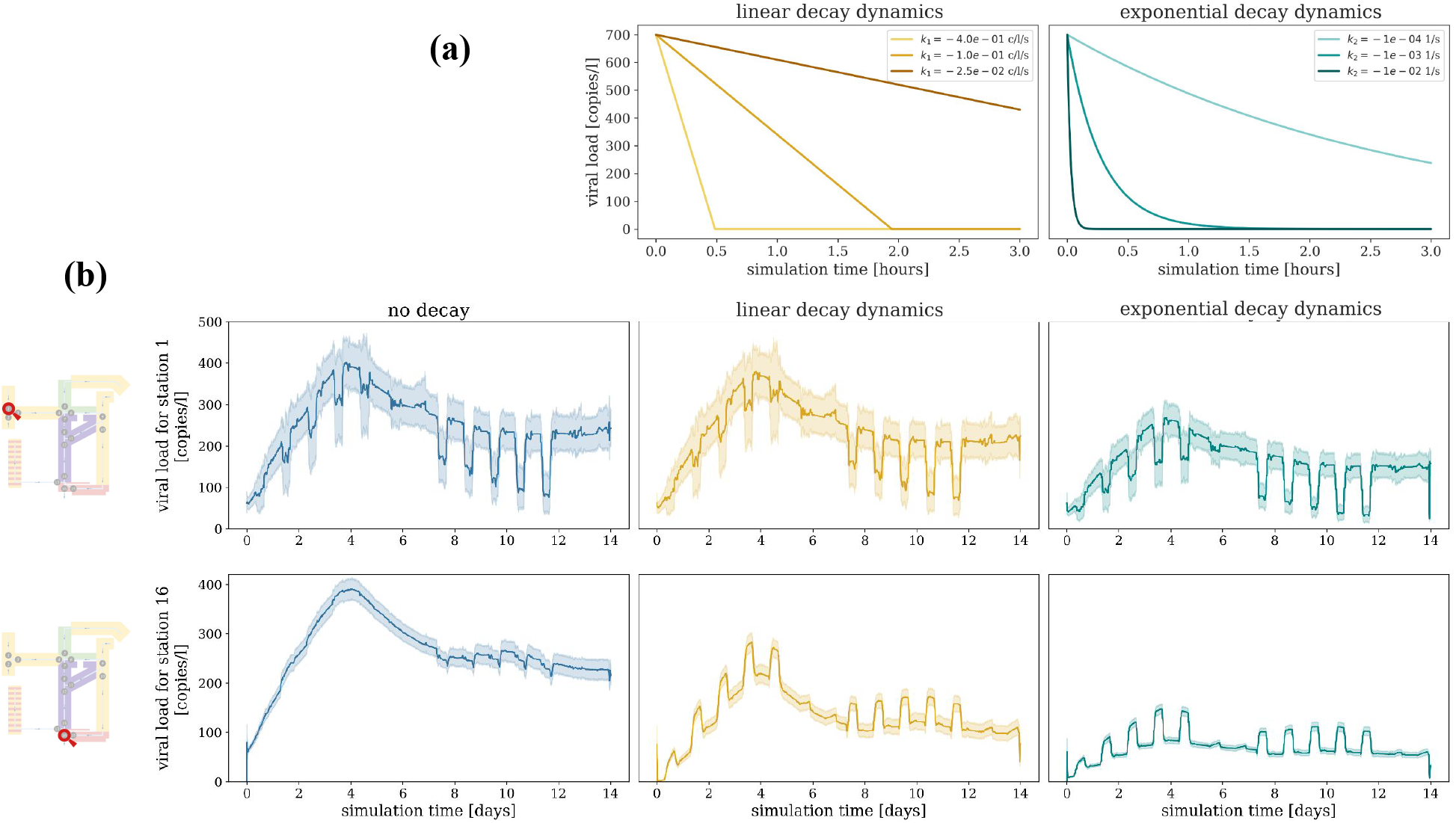
Viral Decay Dynamics. (a) Illustration of the effects of linear decay (left) vs. exponential decay (right) with 3 example parameter settings each on a starting virus concentration of 700 copies per liter. (b) The virus concentration measured at station 1 (top) and station 16 (bottom) over time without rain and with no decay (left), linear decay with 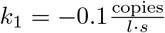 (center), and exponential decay with 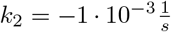 (right).

### 3.7. Virus Normalization

Wastewater systems can have – as outlined above – variable flow rates depending on factors like precipitation and water usage. Hence, wastewater samples have to be normalized to ensure accurate and reliable data interpretation. The two most commonly used normalization strategies are based on either flow rates or additional indicators like the concentration of Pepper Mild Mottle Virus (PMMoV). PMMoV is a plant virus commonly found in human feces at relatively stable concentrations and hence, serves as a good indicator for the amount of human waste in the sample. Here, we use the model to assess which normalization strategy yields corrected viral load values closest to the those one would measure if there was no precipitation event.

The normalization is calculated using one of the following two strategies:

- normalization with flow rates: 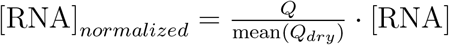
- normalization with PMMoV: 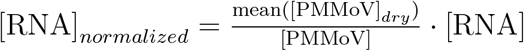

where *Q* is the current flow rate, mean(*Q*_*dry*_) the mean flow rate on dry days, [PMMoV] the current PMMoV concentration, and mean([PMMoV]_*dry*_) the mean concentration of PMMoV on dry days [31]. Rainwater infiltration into a sewer system dilutes PMMoV while increasing flow rates, so correcting wastewater measurements using the ratio between the expected and measured PMMoV concentration or between the measured and expected flow rate can help reduce unwanted variability in wastewater-based data.

We followed exactly these procedures using simulated data. To simulate measurements of PMMoV we assumed a constant PMMoV shedding per agent throughout the simulations and that PMMoV is never subject to viral decay. A small number of observations (between 0.15% and 0.28% per scenario), for which the measured PMMoV concentration was zero copies per liter, were removed. As seen in Fig. 9(b), flow rate normalization (right column) is not effective for correcting wastewater measurements in our model for the effects of either moderate gentle or moderate rain. In both the moderate gentle and moderate rain scenarios, infiltration of rainwater into the model sewer system causes greater proportional increases to the flow rates than decreases to the virus concentrations, so normalization with flow rates leads to notable over-corrections. In contrast, normalization with PMMoV (center column) appears highly effective. Rainwater infiltration affects PMMoV and the virus of interest similarly, so the PMMoV-normalized measurements, while noisier, generally match the norain reference scenario in terms of both shape and scale. Specifically, applying PMMoV normalization reduces the median (across all simulations and time points) of the absolute error between the virus concentration at station 16 for the moderate rain, no decay scenario and the no rain, no decay reference scenario from about 113 to about 8 copies per liter (see Fig. 9(d) and Supplementary Table C.5). Since our PMMoV normalization approach corrects only for rainfall and not for viral decay, PMMoV normalization only marginally improves the median absolute error compared to the reference scenario for the moderate rain, exponential decay and the moderate rain, linear decay scenarios. However, PMMoV appears even less effective when the viral decay is linear rather than exponential. This is likely because in the linear decay scenario, unlike the exponential decay scenario, viral concentrations can and do degrade to zero, which renders normalization useless for certain observations.

**Figure 9:**
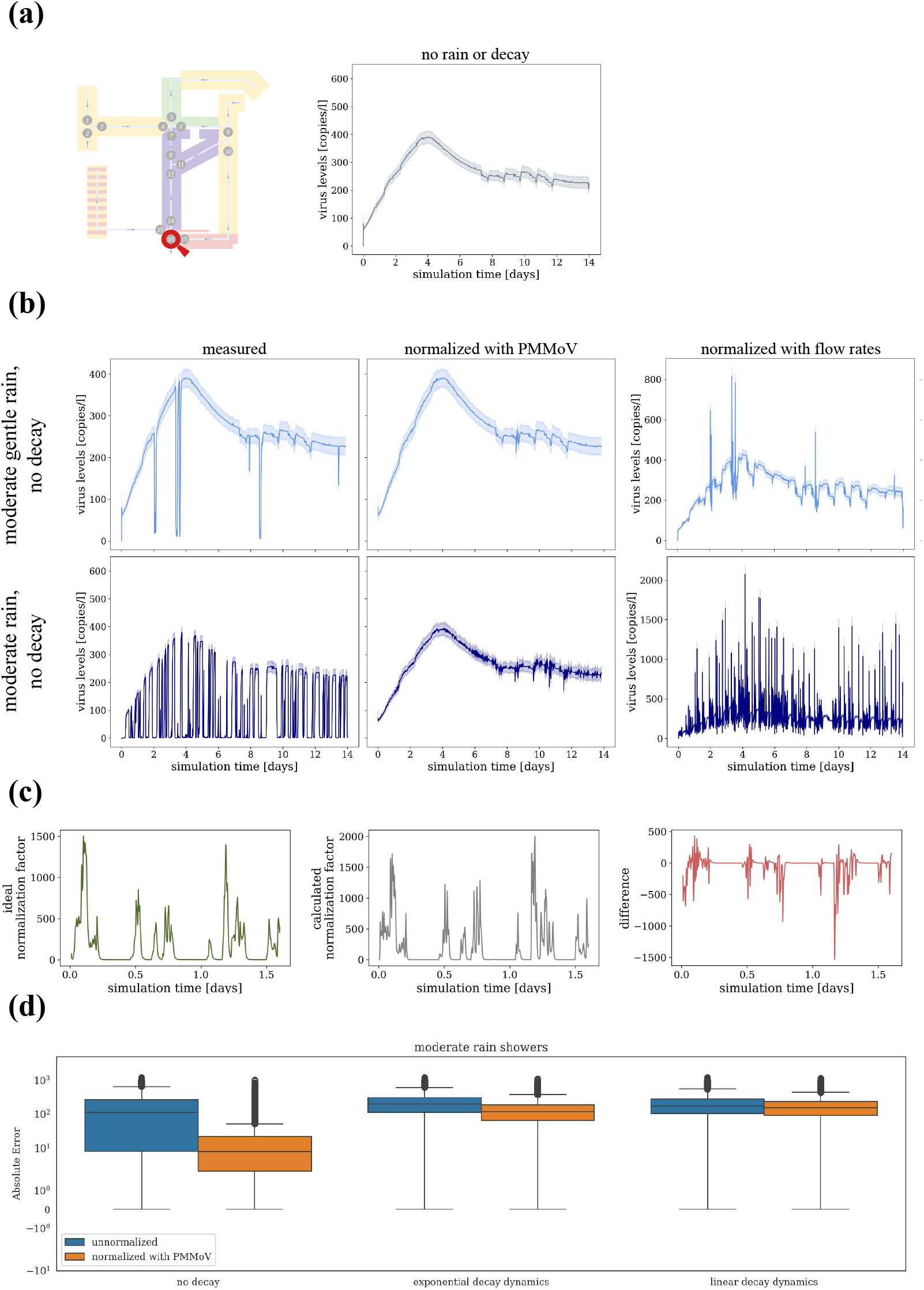
Normalization Strategies Applied to Simulations. (a) The virus concentration at station 16 in the no-rain, no-decay reference scenario. (b) The virus concentration at station 16 in the moderate gentle rain (top) or moderate rain (bottom) scenarios, as measured (left), after normalization with PMMoV (center), and after normalization with flow rates (right). (c) Comparison of the “ideal” normalization factor that would transform the moderate rain results to the no-rain reference results, versus the actual flow-rate-based normalization factor, for one example simulation. (d) The distribution over all simulations and time points of the absolute error between the moderate rain results with no, exponential, or linear decay dynamics and the no-rain, no-decay reference scenario.

Rain reduces the cross-correlations between overall prevalence in the catchment area and the measured RNA copies per liter in wastewater samples over time (see Fig. 10). For the scenarios with rain but without viral decay, applying PMMoV normalization to the wastewater measurements restores the cross-correlation coefficients to very close to their levels in the reference scenario. For the scenarios with both rain and viral decay, PMMoV normalization partially restores the general trends in viral load measurements over time, and therefore has a more limited but non-negligible impact on cross-correlations.

**Figure 10:**
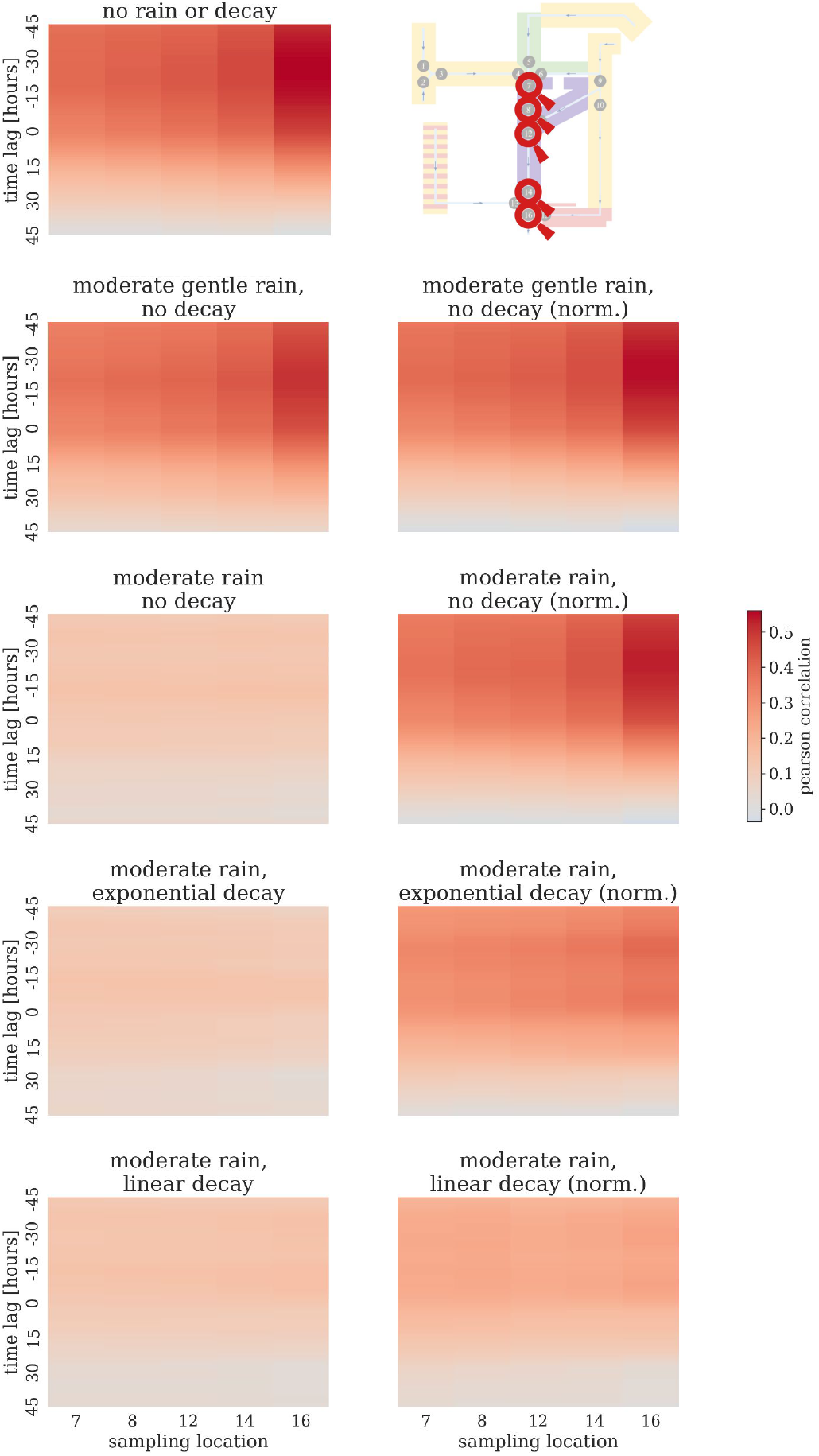
Cross-Correlations Between Wastewater Samples and Prevalence by Scenario. Pearson cross-correlations between RNA copies per liter in wastewater and prevalence over time for five downstream stations, averaged over 250 simulations, for the no-rain, no-decay reference scenario (top left), various decay and rain scenarios without normalization (left column), and the same decay and rain scenarios with PMMoV normalization (right column).

## 4. Discussion

In this study, we presented a first integrative model for the fine-grained description of infectious disease dynamics and wastewater surveillance. The model couples a stochastic model of individual mobility, infection transmission, and disease progression with a highly detailed hydrodynamic model of viral RNA transport through wastewater networks. Using this model for the description of a synthetic neighborhood and corresponding sewer system, we were able to investigate the influence of sampling protocols, precipitation events, virus characteristics, and normalization strategies on the relationship between infection dynamics and resulting wastewater measurements. We found that locations for sampling stations should be chosen carefully, so that they lie downstream of a sufficient number of agents and diverse location types; that precipitation and viral decay can have unexpected, nonlinear impacts on wastewater viral load that require detailed integrative modeling approaches to be understood; and that flow rate normalization should only be implemented with caution as it can lead to large over-corrections if there has recently been precipitation. Overall, our study suggests that if appropriate sampling, normalization, and analysis techniques are used, then wastewater may serve as a leading indicator of disease prevalence.

Sampling station characteristics – including the size of the upstream population, the times of day during which the upstream areas are generally populated, and the distribution of sewer travel times to the station – can both qualitatively and quantitatively affect wastewater measurement dynamics. To produce the highest correlations with prevalence, our study indicates that sampling stations should be placed far enough downstream to receive wastewater from a representative sample of the population of interest as well as from a mix of homes, places of work and study, and recreational areas. If infections are distributed throughout the catchment area, then 24-hour compound sampling can help alleviate some of the disadvantages of upstream sampling location placement, but if infections are localized, downstream placement becomes more crucial, as a sampling station too far upstream may miss the outbreak entirely. However, these considerations must be balanced against others: our study showed how the effects of viral decay become more pronounced the further downstream a sampling station is, and in real-world scenarios, inflow of industrial wastewater may also be a concern. Thus, while we generally argue for midor downstream sampling locations, a detailed modeling approach like ours is needed to choose the optimal sampling location for a particular region of interest.

Our study also illustrates how precipitation and viral decay, separately or together, can have complex and nonlinear impacts on the measured viral load in wastewater, which need to be accounted for when using wastewater data as a public health indicator. Due to the interactions between evaporation, water retention, and other factors, the influence of rainfall on sewer flow rates and viral concentrations within different pipes can be difficult to predict; for example, our model shows how a continuous drizzle can potentially lead to discrete drops in wastewater measurements. Although using sensors to measure sewer flow rates at a sampling station and then adjusting wastewater measurement accordingly is possible, this approach performed sub-optimally in our study and often led to dramatic over-corrections. We find that normalization with a human fecal indicator such as PMMoV may be preferable in sewer systems that have recently been infiltrated by rainwater. This finding is in contrast to the conclusions of Rainey et al. [36], who recommended normalization with flow rates to account for variations in the size of a sewershed’s service population. As Rainey et al. did not consider the effects of precipitation, future work should explore how best to account for both precipitation and population size when normalizing wastewater measurements for comparisons both over time and across sampling locations.

Like precipitation, viral decay in our study led to both qualitative and quantitative changes in the viral load trajectory measured at a particular station over time. Our decay scenarios were intentionally exaggerated – we considered exponential decay with a 90% reduction time of about 0.6 hours, whereas the actual 90% reduction time of SARS-CoV-2 in wastewater is temperature-dependent but usually estimated to be upwards of five days – but this extreme case illustrates how the signal from surface areas far from the sampling location can be lost if viral decay is not appropriately accounted for. In our study, we found that even in the presence of rapid viral decay, PMMoV normalization could restore non-negligible correlations between prevalence and wastewater measurements, but it could only slightly reduce the absolute error relative to a no-rain, no-decay reference scenario. Since PMMoV is very stable in wastewater, normalization with PMMoV alone cannot account for the effects of viral decay on wastewater measurements. Instead, an appropriate viral decay model should be chosen based on the characteristics of the virus of interest and the sewer system, including temperature [14] and biofilms [44] – both of which are likely to be affected in turn by the amount of precipitation in the pipes. Due to its level of detail, our study provides new insights into the interactions between rain, viral decay, and wastewater measurements and underscores the importance of appropriate normalization and analysis of wastewater data.

Overall, our study indicates that despite potential confounding factors, if appropriate sampling, normalization, and analysis techniques are utilized, then wastewater-based surveillance data can provide insights into trends in disease prevalence and possibly predict outbreak peaks 1 to 2 days before they occur. Viral load measurements in wastewater depend not only on the total number of people shedding, but also on the viral load – and, by extension, the infectiousness – of each prevalent infection. High viral loads in wastewater indicate high infection potential across the catchment area, meaning that new infections are likely to occur soon; thus, in our model, the peak in viral load measurements at the furthest downstream station tended to occur about 30 hours before the corresponding peak in overall prevalence. Previous studies, such as the one by Peccia et al. [34], have found that epidemiological measures such as positive test counts and hospital admissions tend to lag several days behind wastewater measurements, and our study suggests that this may not be entirely due to reporting delays. Thus, our results support Peccia et al.’s conclusion that wastewater-based surveillance data can help guide public health officials in deciding when to implement or ease infection control measures.

### 4.1. Limitations and Future Work

One limitation of our model is that we assumed each agent’s water usage to be distributed uniformly across the day. In reality, an individual would only produce wastewater – and, if infected, shed into the sewer system – at certain time points, e.g. while using the bathroom at home after waking up in the morning. We expect that realistic patterns of water usage over time would likely create additional daily trends in wastewater measurements and highlight the advantages of 24-hour compound sampling over grab samples, but future work on this model should incorporate the circadian rhythm of water consumption and shedding behavior to confirm this.

We also leave for future work a sensitivity analysis of how relaxing the assumptions underlying our shedding model might affect our results. Reinfections, vaccinations, and cross-talk between co-circulating pathogens were out of the scope of this initial model, and we therefore did not consider, for example, how the effects of vaccinations on viral shedding [11] could complicate the relationship between disease dynamics and wastewater measurements. Due to data limitations, we assumed that viral shedding in urine and feces was proportional to respiratory shedding and that the peak viral load value was the same for all symptomatic infections, although realistic variations in this value across individuals might decrease the cross-correlations between prevalence and wastewater measurements when the number of infections is small.

Finally, the methods we have so far only applied in the context of an synthetic neighborhood should, in the future, be adapted to real-world scenarios. This will require, for example, introducing an appropriate noise model to account for the effects of wastewater measurement uncertainty and detection limits. Thus, our model’s potential ability to map real-world wastewater measurements back to underlying prevalence remains to be tested.

### 4.2. Conclusions

Our study illustrates the value of sophisticated models of infection and wastewater dynamics and highlights the potential of wastewater-based surveillance data to reflect trends in prevalence without being influenced by sampling bias, reporting delays, or under-ascertainment. While applications to real-world data remain for future work, our simulation study provided key insights into the advantages of downstream sampling location placement, 24-hour compound sampling, models for the effects of viral decay, and PMMoV normalization.

## Data Availability

The version of MEmilio used in this study, including all input files, is publicly available at https://github.com/SciCompMod/memilio/tree/inside-demonstrator-final. Access to the software ++SYSTEMS and its documentation is available upon reasonable request to; all parameter files and simulation outputs used in this study are available at https://doi.org/10.5281/zenodo.14046493. Results were analyzed using the Python code available at https://github.com/inside-consortium/inside demonstrator.

https://doi.org/10.5281/zenodo.14046493

https://github.com/inside-consortium/inside_demonstrator

## Author Contributions

N.S. – conceptualization, methodology, formal analysis, investigation, writing — original draft, writing — review & editing, visualization; J.B. – methodology, software, writing — original draft, writing — review & editing; A.F.H. – methodology, software, writing — original draft, writing — review & editing, funding acquisition; K.W.S – formal analysis, writing — original draft, writing — review & editing, visualization; D.K. – methodology, software, writing — review & editing; A.W. – validation, writing — review & editing, supervision, funding acquisition; M.J.K – validation, writing — review & editing, supervision, funding acquisition; J.H – validation, writing — review & editing, supervision, project administration, funding acquisition.

## Implementation and Availability

The version of MEmilio used in this study, including all input files, is publicly available at https://github.com/SciCompMod/memilio/tree/inside-demonstrator-final. Access to the software ++SYSTEMS and its documentation is available upon reasonable request to; all parameter files and simulation outputs used in this study are available at https://doi.org/10.5281/zenodo.14046493. Results were analyzed using the Python code available at https://github.com/inside-consortium/insidedemonstrator.

