## Supplement for "Integrative Modeling of the Spread of Serious Infectious Diseases and Corresponding Wastewater Dynamics"

---

### Appendix A. Supplementary Mobility Model

Here, we provide the values for the location types $T^{(i)}_{from}$, $T^{(i)}_{to}$; the values for the location transition probabilities $p_i$; and the formulas for the binary-valued functions $\delta^{(i)}(t, \alpha)$ for every mobility rule $m_i$ from Section 2.2.1 in the main manuscript. The mobility rules used for this study are

1. Go to school
2. Return from school
3. Go to work
4. Return from work
5. Go to shop
6. Return from shop

7. Go to recreation
8. Return from recreation
9. Go to hospital
10. Go to ICU
11. Return home when recovered
12. Get buried

The mobility rules are executed at discrete time points $t_0, ..., t_{max}$ with $t_{k+1} = t_k + \Delta t$. Hence, for every $t \in [t_0, t_{max}]$, there are time points $t_k$ and $t_{k+1}$ such that $t \in [t_k, t_{k+1})$. The time is modeled in steps of hours, i.e. $\Delta t = 1$ [hour]. A description of all parameters used in the mobility rules and their values is given in Table C.3.

1. Go to school

$$\begin{aligned} T^{(1)}_{from} &= Home \\ T^{(1)}_{to} &= School \\ p_1 &= SchoolRatio(\mathcal{A}^{(\alpha)}) \\ \delta^{(1)}(t,\alpha) &= \begin{cases} 1, & \text{if } t^{(\alpha)}_{ToSchool} \in \left[t_k - 24 \cdot \lfloor \frac{t_k}{24} \rfloor ; t_{k+1} - 24 \cdot \lfloor \frac{t_{k+1}}{24} \rfloor \right) \wedge \lfloor \frac{t}{24} \rfloor (\bmod 7) < 5 \\ 0, & \text{else} \end{cases} \end{aligned}$$

$t^{(\alpha)}_{ToSchool} \sim U\left(MinSchoolHour, MaxSchoolHour\right)$ is the day time in hours agent $\alpha$ goes to school.

2. Return from school

$$\begin{aligned} T^{(2)}_{from} &= School \\ T^{(2)}_{to} &= Home \\ p_2 &= 1 \\ \delta^{(2)}(t,\alpha) &= \begin{cases} 1, & \text{if } t^{(\alpha)}_{FromSchool} \in \left[t_k - 24 \cdot \lfloor \frac{t_k}{24} \rfloor ; t_{k+1} - 24 \cdot \lfloor \frac{t_{k+1}}{24} \rfloor \right) \\ 0, & \text{else} \end{cases} \end{aligned}$$

$t^{(\alpha)}_{FromSchool} \sim U\left(MinReturnSchoolHour, MaxReturnSchoolHour\right)$ is the day time in hours agent $\alpha$ returns from school.

3. Go to work

$$\begin{aligned} T^{(3)}_{from} &= Home \\ T^{(3)}_{to} &= Work \\ p_3 &= WorkRatio(\mathcal{A}^{(\alpha)}) \\ \delta^{(3)}(t,\alpha) &= \begin{cases} 1, & \text{if } t^{(\alpha)}_{ToWork} \in \left[t_k - 24 \cdot \lfloor \frac{t_k}{24} \rfloor ; t_{k+1} - 24 \cdot \lfloor \frac{t_{k+1}}{24} \rfloor \right) \wedge \lfloor \frac{t}{24} \rfloor \pmod 7 < 5 \\ 0, & \text{else} \end{cases} \end{aligned}$$

$t^{(\alpha)}_{ToWork} \sim U\left(MinWorkHour, MaxWorkHour\right)$ is the day time in hours agent $\alpha$ goes to work.

4. Return from work

$$\begin{aligned} T^{(4)}_{from} &= Work \\ T^{(4)}_{to} &= Home \\ p_4 &= 1 \\ \delta^{(4)}(t,\alpha) &= \begin{cases} 1, & \text{if } t^{(\alpha)}_{FromWork} \in \left[t_k - 24 \cdot \lfloor \frac{t_k}{24} \rfloor ; t_{k+1} - 24 \cdot \lfloor \frac{t_{k+1}}{24} \rfloor \right) \\ 0, & \text{else} \end{cases} \end{aligned}$$

$t^{(\alpha)}_{FromWork} \sim U\left(MinReturnWorkHour, MaxReturnWorkHour\right)$ is the day time in hours agent $\alpha$ returns from work.

5. Go to shop

$$\begin{aligned} T^{(5)}_{from} &= Home \\ T^{(5)}_{to} &= Shop \\ p_5 &= 1 - \exp\left(-\Delta t \cdot ShopRate(\mathcal{A}^{(\alpha)})\right) \\ \delta^{(5)}(t,\alpha) &= \begin{cases} 1, & \text{if } \lfloor \frac{t}{24} \rfloor \pmod 7 < 6 \wedge t - 24 \cdot \lfloor \frac{t}{24} \rfloor \in (7; 22) \\ 0, & \text{else} \end{cases} \end{aligned}$$

6. Return from shop

$$\begin{aligned} T^{(6)}_{from} &= Shop \\ T^{(6)}_{to} &= Home \\ p_6 &= 1 \\ \delta^{(6)}(t,\alpha) &= \begin{cases} 1, & \text{if } \tau^{(\alpha)}_{loc} \geq 1 \\ 0, & \text{else} \end{cases} \end{aligned}$$

$\tau^{(\alpha)}_{loc}$ is the time in hours agent $\alpha$ is at its current location.

7. Go to recreation

$$
\begin{aligned}
T_{from}^{(7)} &= Home \\
T_{to}^{(7)} &= Recreation \\
p_7 &= 1 - \exp\left(-\Delta t \cdot RecreationRate(\mathcal{A}^{(\alpha)})\right) \\
\delta^{(7)}(t,\alpha) &= \begin{cases} 1, & \text{if } \left(\lfloor \frac{t}{24} \rfloor (\bmod 7) < 5 \wedge t - 24 \cdot \lfloor \frac{t}{24} \rfloor \in [19;22)\right) \vee \\ & \quad \left(\lfloor \frac{t}{24} \rfloor (\bmod 7) \geq 5 \wedge t - 24 \cdot \lfloor \frac{t}{24} \rfloor \in [10;22)\right) \\ 0, & \text{else} \end{cases}
\end{aligned}
$$

8. Return from recreation

$$
\begin{aligned}
T_{from}^{(8)} &= Recreation \\
T_{to}^{(8)} &= Home \\
p_8 &= 1 \\
\delta^{(8)}(t,\alpha) &= \begin{cases} 1, & \text{if } \tau_{loc}^{(\alpha)} \geq 2 \wedge t - 24 \cdot \lfloor \frac{t}{24} \rfloor \geq 20 \\ 0, & \text{else} \end{cases}
\end{aligned}
$$

9. Go to hospital

$$
\begin{aligned}
T_{from}^{(9)} &= \{Home, School, Work, Shop, Recreation\} \\
T_{to}^{(9)} &= Hospital \\
p_9 &= 1 \\
\delta^{(9)}(t,\alpha) &= \begin{cases} 1, & \text{if } s^{(\alpha)}(t) = I_{sev} \\ 0, & \text{else} \end{cases}
\end{aligned}
$$

10. Go to ICU

$$
\begin{aligned}
T_{from}^{(10)} &= \{Home, School, Work, Shop, Recreation, Hospital\} \\
T_{to}^{(10)} &= ICU \\
p_{10} &= 1 \\
\delta^{(10)}(t,\alpha) &= \begin{cases} 1, & \text{if } s^{(\alpha)}(t) = I_{cri} \\ 0, & \text{else} \end{cases}
\end{aligned}
$$

11. Return home when recovered

$$T^{(11)}_{from} = \{Hospital, ICU\}$$
$$T^{(i)}_{to} = Home$$
$$p_{11} = 1$$
$$\delta^{(11)}(t, \alpha) = \begin{cases} 1, & \text{if } s^{(\alpha)}(t) = R \\ 0, & \text{else} \end{cases}$$

12. Get buried

$$T^{(12)}_{from} = ICU$$
$$T^{(12)}_{to} = Cemetery$$
$$p_{12} = 1$$
$$\delta^{(12)}(t, \alpha) = \begin{cases} 1, & \text{if } s^{(\alpha)}(t) = D \\ 0, & \text{else} \end{cases}$$

### Appendix B. Supplementary Figures

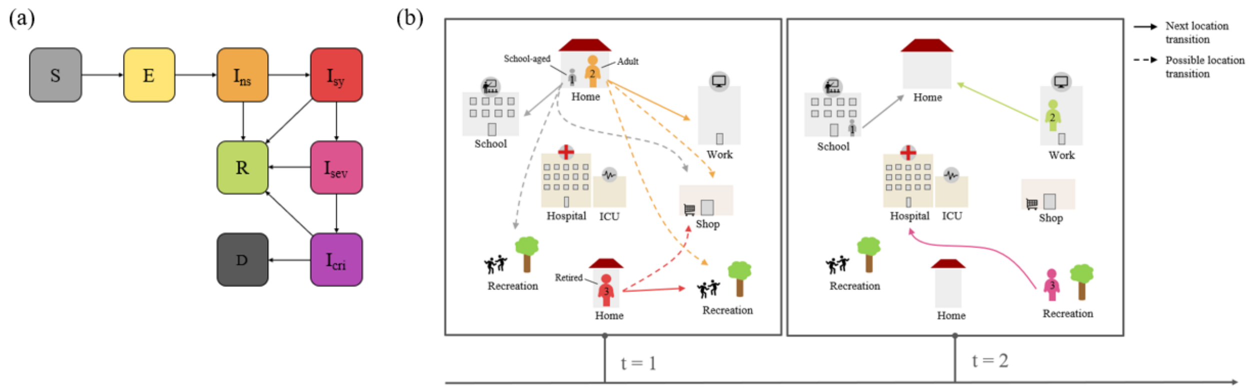


Figure B.1: **Overview of Infection States and Mobility in the ABM.** (a) All potential courses of the disease. The Exposed state always leads to an infectious, non-symptomatic state while for all subsequent infection states (except Recovered or Dead) recovery or worsening of symptoms is possible. (b) Possible location transitions for three agents. The next location transition depends on the agent – i.e. its current location, its age group and infection state – and on time.

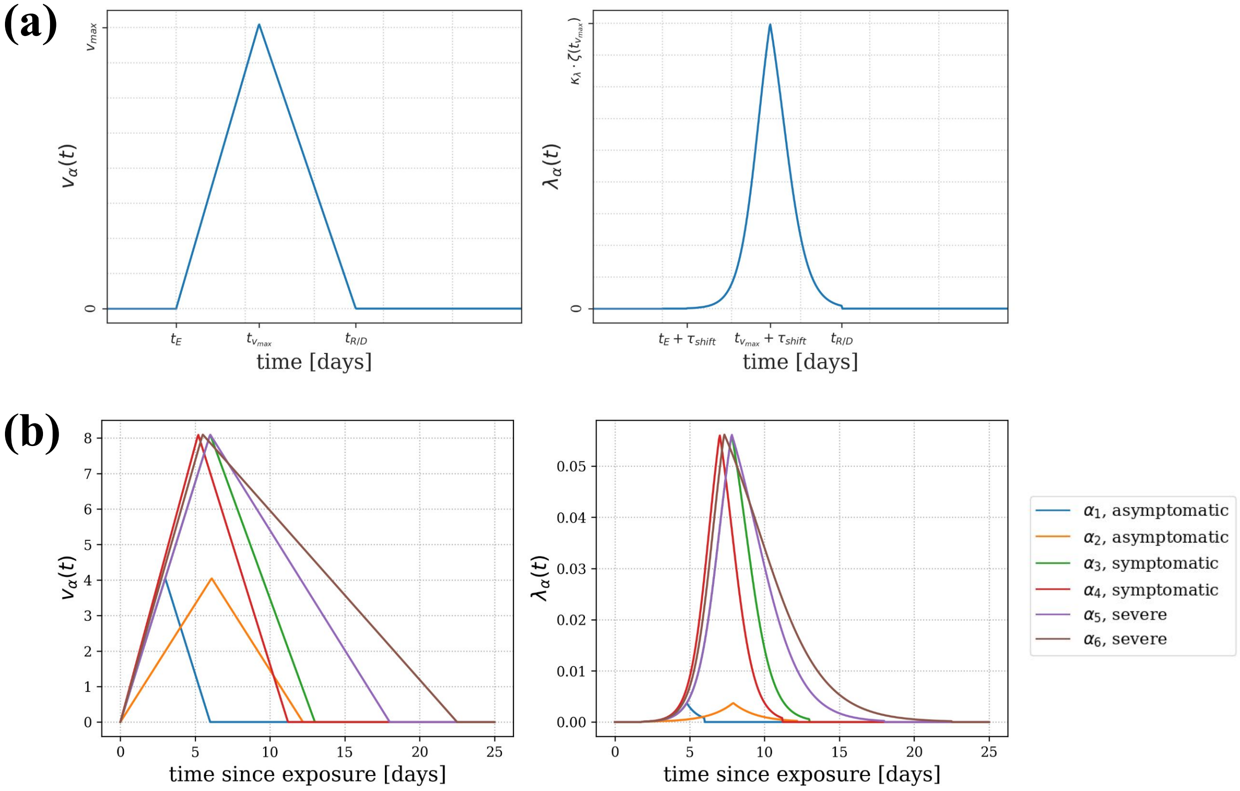


Figure B.2: **Viral Load and Infectiousness Curve.** The (logarithmic) viral load $v_\alpha(t)$ (left) and the infectiousness curve $\lambda_\alpha(t)$ (right) are shown for a generic agent (a) and for six example agents with differing courses of infection (b). $v_\alpha(t)$ is given on the $log_{10}$ scale.

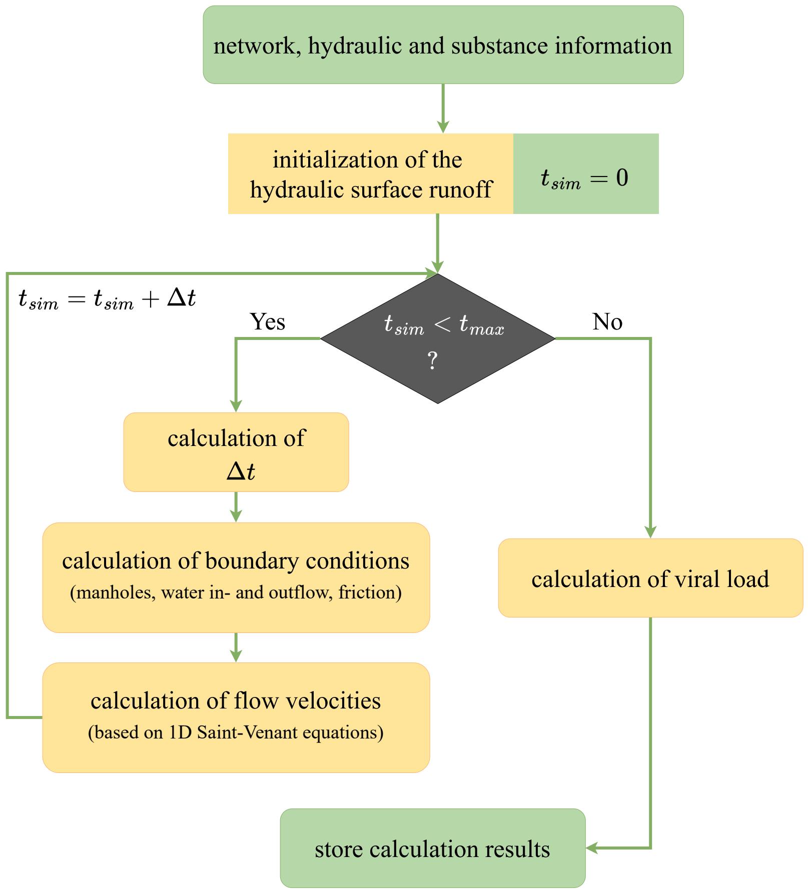


Figure B.3: **Pipeline of Hydrodynamic Calculations.** Based on detailed sewage network information, the hydraulic initial and boundary conditions, and information of the substance of interest, the hydraulic runoff is initialized for the simulation start $t_{sim} = 0$. Iteratively, the time step of the hydraulic calculations $\Delta t$, new boundary conditions, and the corresponding flow velocities of the time step are calculated until $t_{sim}$ reaches the simulation end time point $t_{max}$. Based on the calculated flow rates, the viral load is simulated over time and per location before all simulation results are stored.

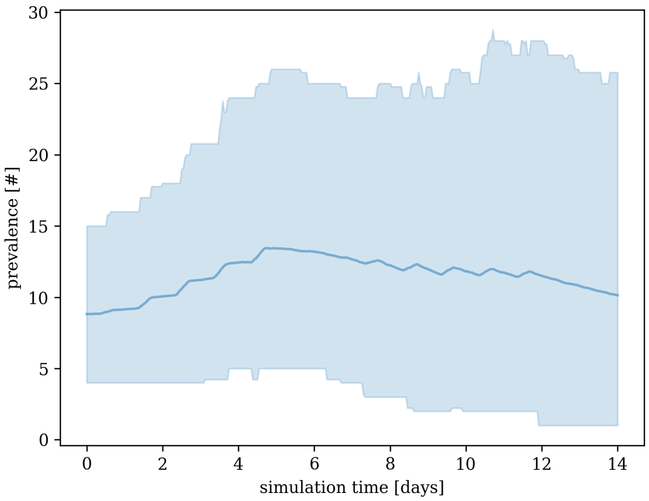


Figure B.4: **Prevalence Simulated by the ABM.** The mean prevalence $(E + I_{ns} + I_{sy} + I_{sev} + I_{cri})$ over time of 250 simulations is shown by a solid line; 95% percentiles are shown by the shaded area.

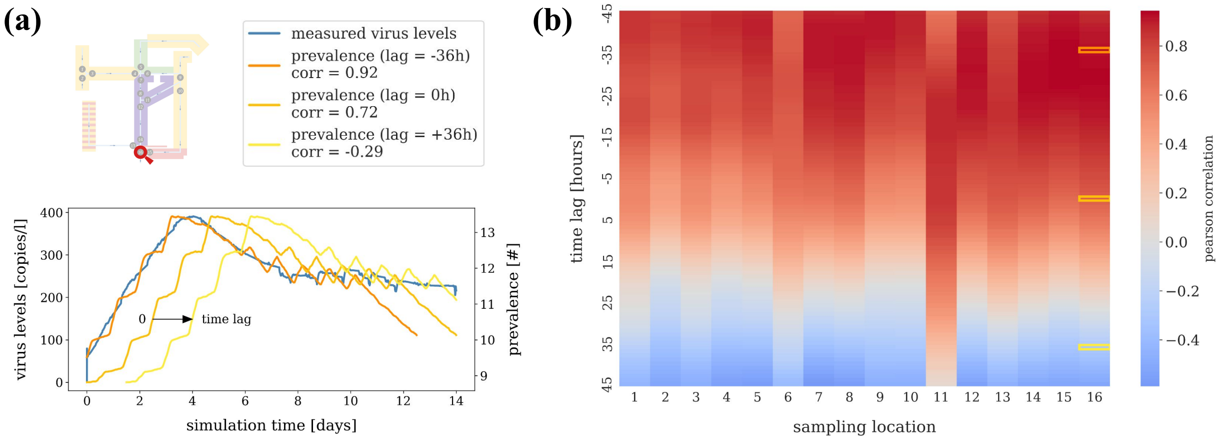


Figure B.5: **Cross-Correlations Between Mean Wastewater Samples and Prevalence.** (a) Trajectory of the mean RNA copies per liter in wastewater (averaged across 250 simulations) for sampling location 16 and the corresponding mean total prevalence shifted with lags -36, 0, and 36 hours. (b) Pearson cross-correlations between the mean RNA copies per liter in wastewater (averaged across 250 simulations) and the corresponding mean prevalence over time for the 16 different locations. The time lag describes the shift in prevalence.

### Appendix C. Supplementary Tables

| Parameter | Description | Default value (per age group) |
|---|---|---|
| $\mu_{I_{ns}}^{I_{sy}}$ | Proportion of symptomatic cases per non-symptomatic case | 0.79 |
| $\mu_{I_{sy}}^{I_{sev}}$ | Proportion of severe cases per symptomatic case | 0.08 |
| $\mu_{I_{sev}}^{I_{cri}}$ | Proportion of critical cases per severe case | 0.18 |
| $\mu_{I_{cri}}^{D}$ | Proportion of dead cases per critical case | 0.1 |
| $\tau_E$ | Duration (in days) of exposed, non-infectious state before transition to non-symptomatic infectious state. Is log-normally distributed. | mean: 3; sd: 1.2 |
| $\tau_{I_{ns}}^{I_{sy}}$ | Duration (in days) of non-symptomatic infectious state before transition to symptomatic infectious state. Is log-normally distributed. | mean: 2.2; sd: 0.5 |
| $\tau_{I_{ns}}^{R}$ | Duration (in days) of non-symptomatic infectious state before recovery. Is log-normally distributed. | mean: 9.2; sd: 2 |
| $\tau_{I_{sy}}^{I_{sev}}$ | Duration (in days) of symptomatic infectious state before transition to severe infectious state. Is log-normally distributed. | mean: 10.5 (1-3), 6 (4-6); sd: 1.1 |
| $\tau_{I_{sy}}^{R}$ | Duration (in days) of symptomatic infectious state before recovery. Is log-normally distributed. | mean: 7; sd: 2 |
| $\tau_{I_{sev}}^{I_{cri}}$ | Duration (in days) of severe infectious state before transition to critical infectious state. Is log-normally distributed. | mean: 5; sd: 2 |
| $\tau_{I_{sev}}^{R}$ | Duration (in days) of severe infectious state before recovery. Is log-normally distributed. | mean: 5 (1-2), 6 (3), 8 (4), 10 (5), 12 (6); sd: 2 (1-5), 3 (6) |
| $\tau_{I_{cri}}^{D}$ | Duration (in days) of critical infectious state before death. Is log-normally distributed. | mean: 6 (1-3), 16.5 (4-5), 11 (6); sd: 2 |
| $\tau_{I_{cri}}^{R}$ | Duration (in days) of critical infectious state before recovery. Is log-normally distributed. | mean: 7 (1-3), 17.5 (4-5), 12.5 (6); sd: 3 |
| $\tau_{infected}$ | Total duration of infection (in days) for a particular agent, i.e. $\tau_E + \tau_{I_{ns}}^{I_{sy}} + \tau_{I_{sy}}^{I_{sev}} + \tau_{I_{sev}}^{I_{cri}} + \tau_{I_{cri}}^{R}$ | |
| $n_A$ | Number of age groups | 6 |

Table C.1: **ABM transmission parameters.**

| Parameter | Description | Default Value |
|---|---|---|
| $v^{sy}_{\max}$ | Peak viral load value in $log_{10}$ units for symptomatic infections | 8.1 |
| $a$ | The viral load value in $log_{10}$ units at which the sigmoid function $\zeta$ of an agent's viral load changes from concave up to concave down | -7.0 |
| $b$ | The slope of the sigmoid function $\zeta$ relative to the change in viral load at the boundary determined by $a$ | 1.0 |
| $\kappa_\lambda$ | Scaling factor applied to the sigmoid function $\zeta$ of an agent's viral load to transform it to a transmission rate | 0.075 |
| $\kappa_\gamma$ | Scaling factor (in RNA copies per day) applied to the sigmoid function $\zeta$ of an agent's viral load to transform it to an RNA shedding rate | $10^{7.1}$ |

Table C.2: **Shedding and Infectiousness Curve Parameters.**

| Parameter | Description | Default value (per age group) |
|---|---|---|
| $T^{(i)}_{from}$ | Start location type for mobility rule $m_i$ | |
| $T^{(i)}_{to}$ | End location type for mobility rule $m_i$ | |
| $SchoolRatio(\mathcal{A})$ | Proportion of persons in age group $\mathcal{A}$ that go to school | 0 (1, 3-6), 1 (2) |
| $MinSchoolHour$ | Earliest day time (in hours) a person can go to school | 6 |
| $MaxSchoolHour$ | Latest day time (in hours) a person can go to school | 9 |
| $t^{(\alpha)}_{ToSchool}$ | Day time (in hours) agent $\alpha$ goes to school | Uniformly distributed in $MinSchoolHour$ and $MaxSchoolHour$ |
| $MinReturnSchoolHour$ | Earliest day time (in hours) a person can return from school | 14 |
| $MaxReturnSchoolHour$ | Latest day time (in hours) a person can return from school | 17 |
| $t^{(\alpha)}_{FromSchool}$ | Day time (in hours) agent $\alpha$ returns from school | Uniformly distributed in $MinReturnSchoolHour$ and $MaxReturnSchoolHour$ |
| $WorkRatio(\mathcal{A})$ | Proportion of persons in age group $\mathcal{A}$ that go to work | 0 (1-2, 5-6), 1 (3-4) |
| $MinWorkHour$ | Earliest day time (in hours) a person can go to work | 6 |
| $MaxWorkHour$ | Latest day time (in hours) a person can go to work | 9 |
| $t^{(\alpha)}_{ToWork}$ | Day time (in hours) agent $\alpha$ goes to work | Uniformly distributed in $MinWorkHour$ and $MaxWorkHour$ |
| $MinReturnWorkHour$ | Earliest day time (in hours) a person can return from work | 15 |
| $MaxReturnWorkHour$ | Latest day time (in hours) a person can return from work | 18 |
| $t^{(\alpha)}_{FromWork}$ | Day time (in hours) agent $\alpha$ returns from work | Uniformly distributed in $MinReturnWorkHour$ and $MaxReturnWorkHour$ |
| $ShopRate(\mathcal{A})$ | Parameter for exponential distribution to decide whether a person of age $\mathcal{A}$ goes to a shop | 1 |
| $RecreationRate(\mathcal{A})$ | Parameter for exponential distribution to decide whether a person of age $\mathcal{A}$ goes to a recreation location | 1 |
| $\tau^{\alpha}_{loc}$ | Duration (in hours) agent $\alpha$ is at its current location $l^{(\alpha)}$ | |

Table C.3: **ABM Mobility Parameters.**

| Id | Rain Scenario | Viral Degradation | Post-Processing |
|---|---|---|---|
| 1a | No precipitation | None | None |
| 1b | No precipitation | None | 24-hour compound sampling |
| 1c | No precipitation | None | Daily grab sampling |
| 2a | Moderate gentle rain | None | None |
| 2b | Moderate gentle rain | None | PMMoV normalization |
| 2c | Moderate gentle rain | None | Flow rate normalization |
| 3a | Moderate rain | None | None |
| 3b | Moderate rain | None | PMMoV normalization |
| 3c | Moderate rain | None | Flow rate normalization |
| 4 | No precipitation | linear ($k_1 = -0.1$) | None |
| 5 | No precipitation | exponential ($k_2 = -0.001$) | None |
| 6a | Moderate rain | linear ($k_1 = -0.1$) | None |
| 6b | Moderate rain | linear ($k_1 = -0.1$) | PMMoV normalization |
| 7a | Moderate rain | exponential ($k_2 = -0.001$) | None |
| 7b | Moderate rain | exponential ($k_2 = -0.001$) | PMMoV normalization |

Table C.4: **Overview of Seven Hydraulic Scenarios.** Simulations using the same 250 ABM output simulations and ++SYSTEMS settings. Some hydraulic scenarios were post-processed in multiple forms to analyze different characteristics.

| degradation setting | rain scenario | MAE (normalized) | MAE (unnormalized) | MAE difference |
|---|---|---|---|---|
| no degradation | moderate gentle | 1.39 | 9.09 | -7.71 |
| | moderate | 17.06 | 161.71 | -144.65 |
| exponential degradation | moderate | 140.83 | 228.27 | -87.44 |
| linear degradation | moderate | 178.91 | 210.19 | -31.28 |

Table C.5: **Effect of PMMoV Normalization**. The mean absolute error (MAE) across all simulations and time points relative to the no-rain, no-decay reference scenario, with versus without PMMoV normalization.
